# Childhood trajectories of internalising and externalising problems associated with a polygenic risk score for neuroticism in a UK birth cohort study

**DOI:** 10.1101/2021.12.15.21267397

**Authors:** Ilaria Costantini, Hannah Sallis, Kate Tilling, Daniel Major-Smith, Rebecca M Pearson, Daphne-Zacharenia Kounali

## Abstract

Neuroticism represents a personality disposition towards experiencing negative emotions more frequently and intensely. Longitudinal studies suggest that neuroticism increases risk of several psychological and physical problems. Improved understanding of how this trait manifests in early life could help inform preventative strategies in those liable to neuroticism.

This study explored how a polygenic risk score (PRS) for neuroticism is expressed from infancy to late childhood across various psychological outcomes and how it associates with trajectories of internalising and externalising problems from ages 4-11 in the Avon Longitudinal Study of Parents and Children (N=5,279). We employed multivariable linear and ordinal regression models to estimate associations between a child neuroticism PRS and psychological outcomes. A three-level mixed-effect model was employed to characterise child internalising and externalising trajectories and estimate how a child PRS associated with both their overall levels and rates of change.

We found evidence that the PRS for neuroticism was associated with a more sensitive temperament in early infancy in addition to higher emotional and behavioural problems and a higher risk of being diagnosed with a variety of clinical disorders, particularly anxiety disorders, in childhood. We also found strong evidence that the PRS for neuroticism was associated with overall levels of internalising and externalising trajectories, with a larger magnitude of effect on the internalising trajectory. The PRS was also associated with slower rates of reduction of internalising problems.

Our findings using a large, well-characterised birth cohort study suggest that phenotypic manifestations of a PRS for adult neuroticism can be detected as early as in infancy and that this PRS associates with several mental health problems and differences in emotional trajectories across childhood.

## Introduction

Neuroticism refers to a relatively stable personality trait characterised by an individual’s tendency to experience negative emotions more frequently and intensely ^1^. Longitudinal studies suggest that neuroticism is a risk factor for a variety of psychological and physical problems such as anxiety, depression, adverse cardiovascular events, and substance misuse ^2–4^. Consequently, this personality trait represents an important personal, economical and societal burden ^5^. Because of the relatively stable and dispositional nature of neuroticism, screening for this trait has been suggested as a strategy for the identification of high-risk groups and the deployment of preventative interventions ^6,7^. In addition, recent evidence has suggested that a general neuroticism factor overlaps substantially with both internalising (e.g., anxiety and depression symptoms) and externalising (e.g., hyperactivity and conduct problems) problems in childhood and that shifting the target of intervention to the development of emotional coping strategies and resilience in children may be more effective in preventing later onset of mental and physical health problems ^6,7^.

Recent twin and family-DNA informed studies ^8,9^ have suggested a heritability of neuroticism of approximately 30%, indicating a moderate genetic contribution to this trait. In addition, genome-wide association studies (GWASs) have identified hundreds of independent loci contributing to its aetiology ^10,11^. Thus, it is increasingly possible to construct polygenic risk scores (PRS) as indicators of an individual’s genetic liability for neuroticism ^12^ in populations of European ancestry ^13^. While some longitudinal studies have explored phenotypic manifestations of PRSs for various psychiatric disorders (e.g., schizophrenia^14,15^) and childhood psychopathology (e.g., ADHD^16^) in childhood and adolescence, no studies to date have examined the association between a PRS for neuroticism and phenotypic manifestations of this score in infancy or early childhood. In addition, no studies to date have evaluated the association of a PRS for neuroticism with trajectories of emotional and behavioural problems across any age group (see Akingbuwa^17^ for a meta-analysis of these studies). Exploring phenotypic manifestations of a genetic predisposition to psychological traits like neuroticism in infancy and childhood could help in understanding how early life preventative interventions and screening strategies for subsequent disease could be introduced, indicating optimal time-points for screening and targeting high risk populations. In addition, intervening to minimise the risk associated with early life risk factors for later psychiatric outcomes may be a more effective strategy for reducing the burden from these outcomes than treating them directly as such an approach may be more comprehensive in its downstream effects. This may be particularly important in the context of child psychiatry where psychological symptoms are not as differentiated as in later ages (e.g., middle to late adolescence) and were historically understudied and their prevalence underappreciated ^18–20^.

There were two primary aims of this study. First, we aimed to examine phenotypic manifestations of a PRS for neuroticism across a large number of temperamental traits, psychological problems and clinical disorders in infancy and childhood. Second, we aimed to examine how a PRS for neuroticism associates with childhood trajectories of emotional and behavioural problems where repeated measures were available.

As secondary aims of this study, we attempted to estimate the causal effect of the child PRS on the above-mentioned traits. First, we attempted to estimate the causal effect of these association independent of potential maternal genetic confounding (i.e., by adjusting for a maternal PRS for neuroticism). Second, we investigated the extent to which these associations were driven via the effect of a genetic liability to neuroticism *per se* as opposed to the effect being driven by independent pleiotropic pathways. This was done by using a PRS restricted to SNP_S_ associated with neuroticism at *P*<5 × 10^−8^ and by adjusting for maternal PRS for neuroticism.

Finally, evaluating child emotional and behavioural problems often presents a challenge because of the reliance on maternal reports. Some evidence, albeit conflicting, suggests that maternal ratings of child behavioural and emotional problems can lead to misclassification of child psychopathology as the scores may relate more to psychological problems in the mother herself than to the child ^21–24^. By contrast, recent evidence has highlighted that the magnitude of differential misclassification is often negligible ^23,24^. Where differential misclassification could still be a threat to the validity of our analyses, we systematically employed various sensitivity analyses to explore the direction and magnitude of this potential bias.

## Methods

### Participants

The Avon Longitudinal Study of Parents and Children (ALSPAC), also known as Children of the 90s, is an ongoing prospective population-based birth cohort study. Between 1990 and 1992, 14,541 pregnant women were recruited in Bristol and the surrounding area, previously known as Avon County. The original mothers and partners (Generation 0: ALSPAC-G0) ^25^ and their living children (Generation 1: ALSPAC-G1) ^26^ have been followed-up regularly since recruitment through questionnaires and clinic assessments. When the oldest children were approximately 7 years of age, an attempt was made to bolster the initial sample with eligible cases who had failed to join the study originally), resulting in an additional 913 children being enrolled. The total sample size for analyses using any data collected after the age of seven is therefore 15,454 pregnancies, resulting in 15,589 foetuses. Of these 14,901 were alive at 1 year of age ^27^. This study focuses on the index children (ALSPAC-G1) and on how their PRS for neuroticism is expressed across childhood. Please note that the study website contains details of all the data that is available through a fully searchable data dictionary and variable search tool http://www.bristol.ac.uk/alspac/researchers/our-data/. Ethical approval for the study was obtained from the ALSPAC Law and Ethics Committee and South-West National Health Service (NHS) Research Ethics Committee; informed consent for the use of data collected via questionnaires and clinics was obtained from participants following the recommendations of the ALSPAC Ethics and Law Committee at the time. Consent for biological samples has been collected in accordance with the Human Tissue Act (2004).

### Genetic Data

Genotyped data were available on 7,851 children and 7,826 mothers in the ALSPAC study. Details of genotyping and quality control measures are available in **Supplementary Materials**. PRSs for genetic liability for neuroticism were derived for mothers and children and standardised prior to analysis. These scores were based on publicly available summary statistics from a recent GWAS of neuroticism (N=329,821) which identified 116 independent genetic variants at a genome-wide significance threshold (P<5 × 10^−8^) ^10^.

PRS were calculated using PRSice-2^28^ and restricted to SNPs with a minor allele frequency of >1% and an info score of >0.8. Scores were created by summing the number of risk alleles present for each SNP (0, 1, or 2) weighted by the effect estimates from the original GWAS. Thirteen PRS were derived using P-value thresholds ranging from P<0.5 to P<5 × 10^−8^ (**Supplementary Table 1**), with a greater number of SNPs included as the P-value thresholds became less conservative. Our primary analysis used a PRS based on a threshold of P<0.05, which is the threshold that most optimally captured the trade-off between variance explained for neuroticism liability and added error term ^10^. Using this P-value threshold, the neuroticism polygenic score explained 2.79% of the variance in neuroticism (β=0.19, P=2.65□×□10^−47^) ^10^.

### Outcome Measures

#### Carey Infant Temperament Scales (CTSs) – *6 and 24 months*

Carey Infant Temperament Scales (CTSs) ^29,30^ is a common questionnaire used to assess child’s temperament (examples of items: “Child cries when left playing alone”, “child plays quietly with toys”) which comprises a number of age-appropriate questions relating to different temperament domains. In this analysis, we used the prorated score which was corrected for age at completion of the questionnaire and gestation at delivery. The results are coded so that higher scores indicate a more “difficult” temperament. Further details are reported in **Supplementary Materials** and values for each subscale are reported in **Supplementary Table 2**.

#### Strengths and Difficulties Questionnaire (SDQ) – *4 to 11 years*

The Strengths and Difficulties Questionnaire (SDQ) ^31^ is a brief behavioural screening questionnaire validated for children who are approximately 3-16 years old. It provides a score ranging from 0 to 40, where higher scores indicate a larger number of emotional and behavioural difficulties exhibited by the child. Cronbach’s alpha values for each subscale and the derived internalising and externalising subscales are reported in **Supplementary Table 2**. Further details about the subscales and cross-informants’ correlations are reported in **Supplementary Materials**.

#### Locus of control – *8 years*

Children’s locus of control was assessed using an adaptation of the Children’s Nowicki Strickland Internal External scale (CNSIE) ^32^. This form was administered to ALSPAC children when they were tested at 8 years of age. Items were read out loud by the examiners and the child was asked to respond yes or no. They were reminded of the confidentiality of the test and that there were no correct answers. Higher scores indicate a more external locus of control. In the current sample, the internal reliability as measured with Cronbach alpha was low (0.47) (**Supplementary Table 2)**.

#### Self-Esteem *– 8 years*

Children’s self-worth and scholastic competence were assessed with a 12-item shortened form of Harter’s Self Perception Profile for Children ^33^ at around 8 years of age. In the current sample, internal reliability as measured with Cronbach alpha was satisfactory with values of 0.75, 0.69, and 0.65 for the total score, the scholastic self-esteem score and the global self-esteem score, respectively (**Supplementary Table 2)**.

#### Intelligence Quotient (IQ) *– 8 years*

The Wechsler Intelligence Scale for Children (WISC-III UK) ^34^ was used to assess child’s IQ at 8 years of age (**Supplementary Table 2)**.

#### Axis-I Disorders – *7 and 11 years*

Any axis-I disorders (i.e., clinical conditions such as anxiety and depressive disorders) classified using the Diagnostic and Statistical Manual of Mental Disorders (DSM-IV) ^35^ (yes vs no) and the International Statistical Classification of Diseases and Related Health Problems, Tenth Revision (ICD-10) ^36^ were assessed using maternal and a mix of maternal and teacher reports from the Development and Well-being Assessment (DAWBA) ^37,38^ when children were 7 and 10 years old (**Supplementary Table 3)**. Up to six ordinal bands were derived with a computer algorithm which has been shown to be similar in accuracy to a clinical rating method ^38^. Further details are reported in **Supplementary Materials**.

#### Big Five – *13 years*

The emotional stability subscale (i.e., inverse of neuroticism) of the Big Five personality test ^39^ was used as an outcome in our positive control analysis. Negative and positive controls are usually employed in epidemiological analyses to test the validity of the association examined ^40^. Here, we conducted a positive outcome control analysis to test the validity of the assumption that a PRS for neuroticism was indeed associated with measured neuroticism (i.e., the emotional stability subscale of the Big Five). The Big Five questionnaire was measured as part of the TF2 clinic visit when the child was 13 years old. Higher scores on the emotional stability variable indicate higher emotional stability (i.e., lower neuroticism) (**Supplementary Table 2)**.

#### Covariate Measures

In all the separate linear and ordinal regression models we included age of the child, sex of the child and the first five principal components of genetic ancestry as covariates. Age of the child was not included as a covariate in the models where the CTSs were the outcome as the scores were already adjusted for gestational age and age of the child. These covariates were also used in the mixed effect models. In addition, to support the missing at random (MAR) assumption of these models, we included variables indicating maternal education, maternal social class and maternal age as they were strongly associated with the probability of having missing data.

#### Statistical Analyses

Our exposure (child PRS for neuroticism) was standardised using z-scores to have mean=0 and SD=1, with a higher PRS indicating a higher liability to neuroticism.

To address the first aim of this study (i.e., “to examine the phenotypic manifestation of a PRS for neuroticism across childhood”) we employed separate multivariable linear regression models with robust standard errors (SEs) to estimate coefficients and 95% CIs for the associations between the child PRS for neuroticism and different psychological outcomes (i.e., CTSs, SDQ, Locus of control, Self-Esteem and IQ) adjusted for the above-mentioned covariates. Using robust SEs (i.e., Huber-White sandwich estimators) can help in addressing potential violations of linear regression assumptions such as normality, heteroscedasticity, or observations that exhibit large residuals. Primary analyses were conducted on the complete case dataset. As a positive control analysis, we investigated whether the child PRS for neuroticism was associated with the neuroticism (i.e., inverse of emotional stability) subscale as measured by the Big Five at 13 years of age. Ordinal logistic regressions were used when the outcome variables were not continuous integers (i.e., DAWBA).

To take into account an inflated false discovery rate due to multiple testing, a heuristic Bonferroni adjusted P-value threshold was set at P<0.0014 (0.05/35, where 35 pertains to each unique outcome tested). However, as we have included subscales which are highly correlated with each other, this threshold is likely to be overly conservative. We also investigated the potential for bias in our analyses due to levels of attrition in the outcomes in our analysis sample (from 3,579 up to 6,271 individuals with genetic and phenotypic data). To perform this, we generated 100 imputed datasets using multiple imputation through chained equations (MICE) ^41–43^ (see **Supplementary Methods** for further details).

#### Selected linear mixed effect model

In this study, a three-level mixed effect model (command *xtmixed* in Stata-16 ^44^) with random intercept and slopes was employed to characterise behavioural and emotional trajectories over time and to examine the association of the child PRS with the overall score of emotional and behavioural problems and how these change according to sex, type of scale and reporter of the scale. The levels represent a dependence structure where children’s scores at different measurement occasions (level 1) are nested within the type of scale (internalising or externalising) (level 2) which are nested within an individual child (level 3) (**Figure 1**)^45^. This model allows a separate intercept and slope for each child in each scale and it permits the investigation of the effect of covariates as fixed effects. The model included a random intercept and slopes for the change over time which permitted estimation of individual child variation in score levels, and how these change with time across different scales and reporters. Restricted maximum likelihood (REML) was employed instead of maximum likelihood (ML) for fitting this model. Model adequacy was assessed using information criteria and examining model assumptions, namely homogeneity of variance (with group variance being constant between groups and predictors), linearity and normality of residual variances at different levels.

**Figure 1.**
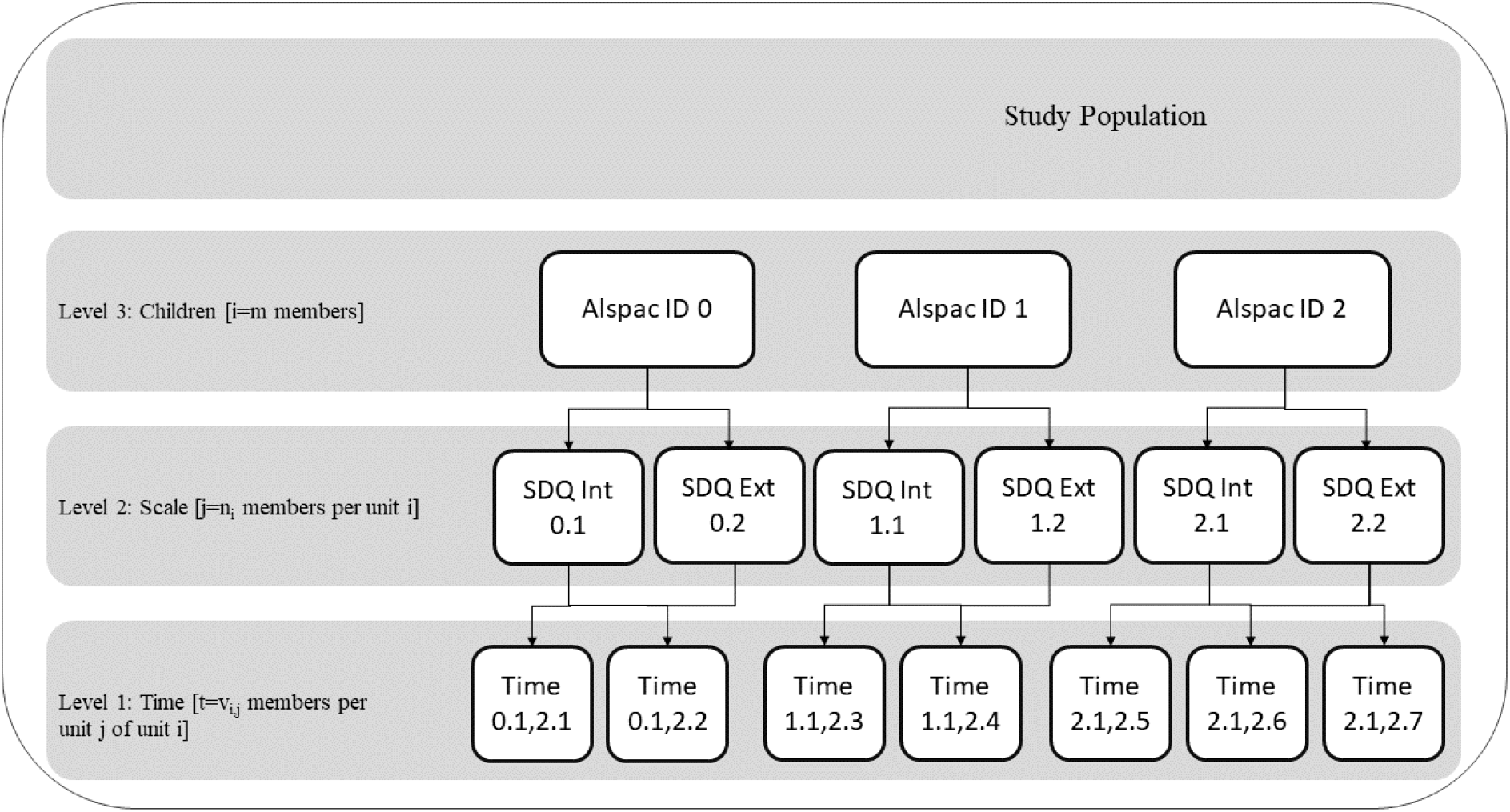
Hierarchical structure of the selected linear mixed effect model. The levels represent a dependence structure where children’s scores at different measurement occasions (level 1) are nested within the type of scale (internalising or externalising) (level 2) which are nested within an individual child (level 3).

The sample used in three level linear mixed models consisted of 5,729 children with at least 1 observation per internalising and externalising subscale over a maximum of 7 occasions per scale. The mean number of measurements was 5.62 (SD=3.50). The majority (74.5%) of the child questionnaires on the SDQ for this study were answered by the mother for the parent reported scale. The median (IQR) age of the assessments where genetic information was available was 8.33 (6.75 – 10.83) years with a minimum age of 3.67 years and a maximum of 14.00 years (age was centred at its median). For these analyses, the natural log (plus 1) values of the outcomes were used for both analytical convenience (e.g., variance stabilisation and ensuring positivity on the outcome scale) as well as ease of interpretation (expressing covariate effects on the ratio scale, i.e., percentage change). Our analyses included all available data on the SDQ scale and the model implies that missing observations over time are missing at random (MAR), conditional on all included covariates. To mitigate the risk of bias due to attrition we included in these models additional covariates that were found to be strongly associated with attrition (i.e., maternal education, maternal social class and maternal age).

We explored the role of the reporter (parent vs teacher) by including it both as a fixed as well as a random effect at the level representing the different scales used for each child. This permitted us to decompose the SDQ score variability further to that associated to different scales used and between reporters. This variance component was additional to between intra-child variability (level 2).

To examine whether the child PRS for neuroticism was associated with the overall level of the trajectories, we included the PRS as a fixed-effect term and we used interaction terms to explore its effects depending on the scale used, the reporter of the scale and the sex of the child. To examine how the PRS was associated with changes over time in our model, we included a main effect of the PRS and an interaction of the PRS with the fixed-effect age term. Code used to perform these analyses and additional information on the model are presented in the **Supplementary Materials**.

#### Sensitivity analyses

We used various sensitivity analyses to test the robustness of our findings. First, we employed different methods to explore the potential presence of differential and dependent measurement error due to the maternal PRS. We used a signed directed acyclic graph (DAG) to illustrate potential measurement error or misclassification as suggested by VanderWeele et al ^46^ (**Supplementary Figure 1**). We firstly used maternal genotype (N=7,826) as the exposure when assessing child’s behaviour as reported by the child’s teacher as an outcome while adjusting for child genotype to test whether maternal own PRS for neuroticism may have influenced teacher’s rating. Second, we developed “difference in score” variables by subtracting the maternally reported score of child’s problems (measured with the SDQ at 8 and 11 years of age) from the teacher reported score of child’s problems (measured with the SDQ at 8 and 11 years of age). Then, we employed linear regression models adjusted for the child PRS to explore the association between the maternal PRS and the difference in score. In addition, to address one of the secondary aims of this study (i.e., “estimating the causal relationship between the child PRS and later emotional and behavioural difficulties”) we adjusted for maternal PRS in the separate multivariable linear regressions in order to account for a possible confounding effect of maternal PRS for neuroticism on child’s outcomes. Finally, to address the last of our secondary aims (i.e., “estimating the causal relationship between child genetic liability to neuroticism and later emotional and behavioural difficulties”) we re-ran separate linear and ordinal regression models using a genome-wide significance threshold (P<5 × 10^−8^) for PRS construction. This could help to disentangle whether the observed effects of the neuroticism PRS are indeed acting through a liability to neuroticism as opposed to alternate pathways. For more details about the sensitivity analyses refer to the **Supplementary Material**’s sensitivity analyses section. All analyses were conducted using Stata 16 statistical software.

## Results

### Descriptive results

The numbers of individuals who participated in the different assessments at various visits are shown in **Supplementary Table 2 and 3**. Of the 7,851 ALSPAC children with genetic data available (51.25% male and 48.75% female), 6,271 to 3,579 participated in assessments from 6 months to 11 years of age, respectively. Of the participants who had genetic information, missingness for the outcomes of interest was modest at 6 months of age (20.42% missingness) and high when the SDQ was measured by the teacher at age 8 (up to 54.50% missingness) (See **Supplementary Figure 2** for a Flow diagram of ALSPAC participants). Participants who had data on the variables included in the different models (e.g., outcome, exposure and covariates) generally had mothers who were of a higher social class, obtained a higher education and who were less likely to have smoked during pregnancy (**Supplementary Table 4**). Correlations between subscales of SDQ and between mother and teacher reports are reported in **Supplementary materials** and **Supplementary Table 5**.

### Association between child PRS for neuroticism and various psychological outcomes

Betas (β) indicate a unit increase or decrease in the outcome score for each 1 SD increase in the exposure. We found strong evidence of an inverse association between child PRS for neuroticism and emotional stability (β_emotional stability_=-0.75, 95% CI: -0.96 to -0.55, P=7.460 × 10^−13^), supporting the internal validity of our exposure.

The earliest behavioural outcome that showed nominal evidence for association with the child PRS was for the approach, adaptability and threshold subscales of the CTS at 6 months of age. Generally, the association between the child PRS for neuroticism and various psychological outcomes strengthened as the age of the children increased. However, evidence to support some of these associations weakened when considering a heuristic P-value threshold for multiple testing (complete findings are presented in **Supplementary Table 6 and Figure 2**).

We found strong evidence that a higher neuroticism PRS was associated with higher scores in both the emotional and behavioural domains of the SDQ (i.e., total score) across all 7 occasions from 4 to 11 years of age. Effect sizes generally increased throughout childhood with the smallest estimates at age 4 (β=0.27, 95% CI: 0.15 to 0.40, P=2.30 × 10^−5^) and the largest at age 8, both when reported by the mother (β=0.52, 95% CI: 0.38 to 0.67, P=2.08 × 10^−12^) and when reported by the teacher (β=0.52, 95% CI: 0.33 to 0.71, P=8.13 × 10^−8^). These findings were consistent independent of whether the outcome was teacher or parent reported (**Supplementary Table 6 and Figure 3**).

In addition, we observed strong evidence of an association between the neuroticism PRS and the likelihood of received clinical diagnoses at both 7 and 10 years of age as measured with the DAWBA. In particular, a higher PRS for neuroticism was associated with a 13% (95% CI: 6 to 20%, P=8.85 × 10^−5^) increased risk of generalised anxiety disorder (GAD) and a 14% (95% CI: 8 to 21%, P=3.61 × 10^−6^) increased risk at 7 and 10 years of age, respectively. Similarly, we found strong evidence that children with a higher PRS for neuroticism were at 13% (95 % CI: 7 to 20%, P=1.90 × 10^−5^) and 19% (95 % CI: 12 to 26%, P=2.80 × 10^−8^) increased risk of any anxiety related disorder at 7 and 10 years of age, respectively. When exploring the association between the PRS for neuroticism and any behavioural disorder as measured as a combined score across three traits (i.e., conduct disorder, oppositional defiant disorder and ADHD), we found nominal evidence that the PRS was associated with a 9% (95% CI: 4 to 16%, P=0.001) and 9% (95% CI: 3 to 16%, P=0.003) increased risk of this combined score at 7 and 10 years of age, respectively. We found little evidence of an association between the child PRS for neuroticism and depressive disorders at 7 years of age (OR=1.05, 95% CI: 0.99 to 1.12, P=0.074), but stronger evidence of an association at 10 years of age (OR=1.11, 95% CI: 1.04 to 1.17, P=0.001) (**Supplementary Table 7 and Figure 4**).

Finally, we also observed strong evidence that a higher PRS for neuroticism was positively associated with broader psychological constructs such as having an external locus of control and inversely associated with global and scholastic self-esteem and IQ (**Supplementary Table 6**).

### Child PRS for neuroticism and trajectories of internalising and externalising problems

The overall mean for the internalising scale of the SDQ was estimated as 2.97 (95% CI: 2.67 to 3.35) for boys when assessed by the parents. In this model, there was evidence for an association between the child PRS for neuroticism and child trajectories of internalising and externalising problems. For every SD increase in child PRS for neuroticism we observed an average 6% (95% CI: 4 to 7%) increase in internalising problems in boys as reported by the parent. The magnitude of this effect was halved (i.e., 3%, 95% CI: 2 to 5%) when examining externalising problems. We also observed strong evidence of a rate of reduction of 3% (95% CI: 2 to 3%) per year of age on the internalising scores in boys when reported by the parents, which was larger than the rate of reduction smaller in girls by 1% (95% CI: 1 to 2%). This rate of reduction was more pronounced on the externalising scale (rising to 6% per year, 95% CI: 4 to 6%). The PRS effect on the rate of change with age became more pronounced on the internalising scale where the rates of change were slower. We observed similar patterns of the effect of the PRS in girls. Differences between sexes mainly consisted of differences in score levels with age according to scale and reporter. Further discussion of findings from this model are reported in the **Supplementary Materials**. Complete findings from fixed and random effects models are presented in **Table 1**. A graphical representation of these marginal model predictions according to age, sex and PRS levels and reporter is presented in **Figures 5 and 6**.

**Table 1.**
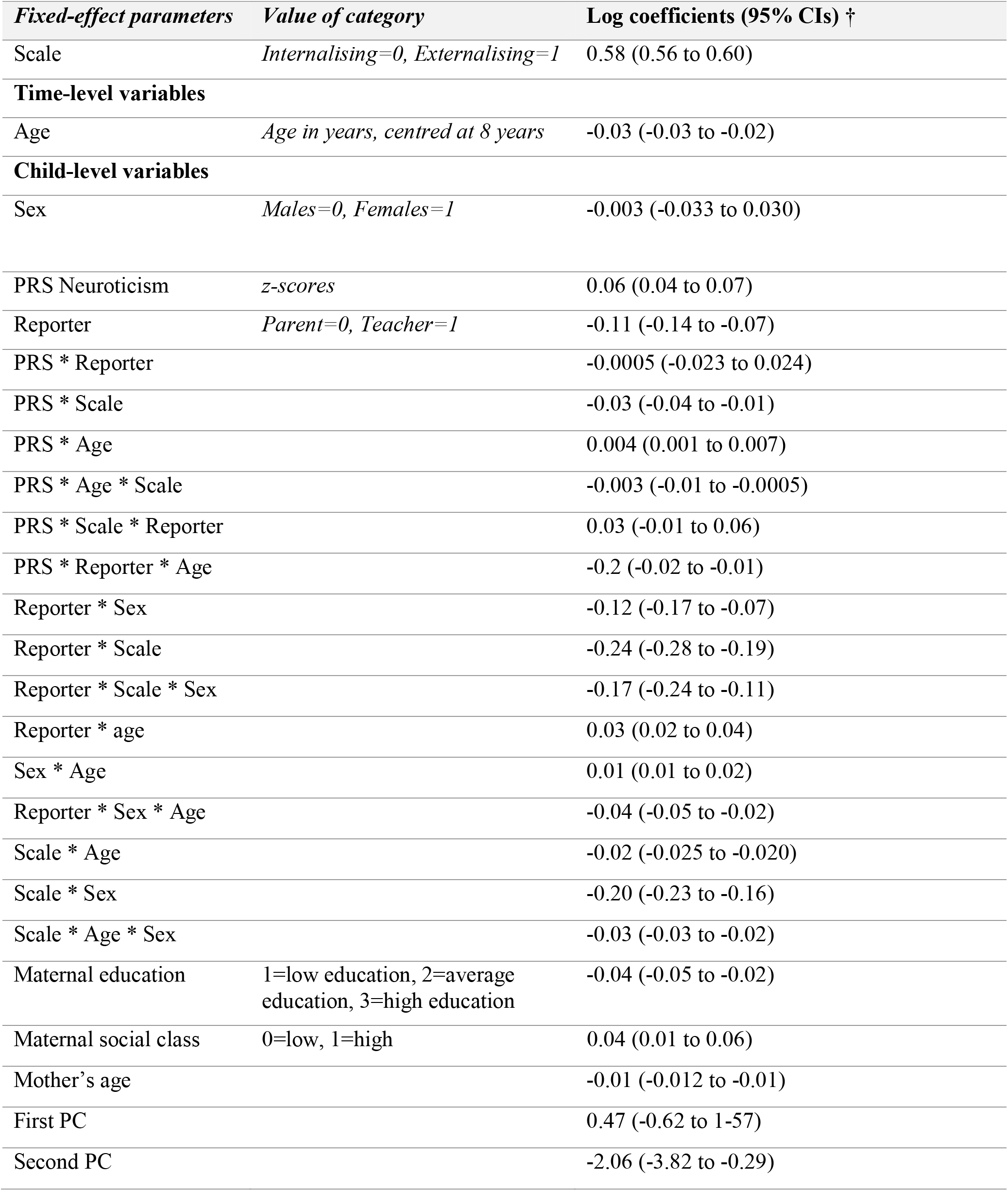
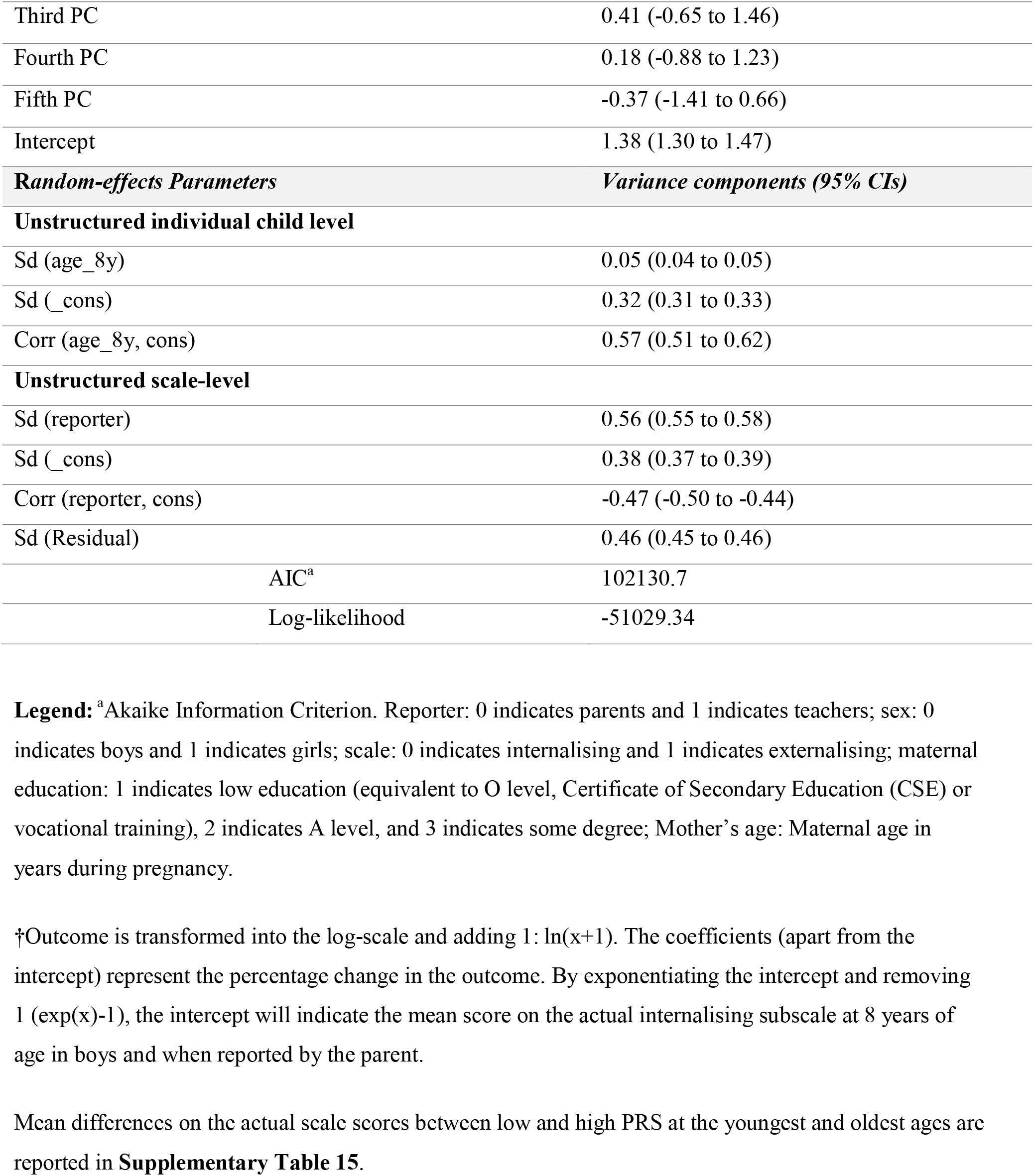
Findings for the three-level model investigating the association between PRS and SDQ across time.

| *Fixed-effect parameters* | *Value of category* | *Log coefficients (95% CIs) †* |
| --- | --- | --- |
| Scale | *Internalising=0, Externalising=1* | 0.58 (0.56 to 0.60) |
| **Time-level variables** |  |  |
| Age | *Age in years, centred at 8 years* | -0.03 (-0.03 to -0.02) |
| **Child-level variables** |  |  |
| Sex | *Males=0, Females=1* | -0.003 (-0.033 to 0.030) |
| PRS Neuroticism | *z-scores* | 0.06 (0.04 to 0.07) |
| Reporter | *Parent=0, Teacher=1* | -0.11 (-0.14 to -0.07) |
| PRS * Reporter |  | -0.0005 (-0.023 to 0.024) |
| PRS * Scale |  | -0.03 (-0.04 to -0.01) |
| PRS * Age |  | 0.004 (0.001 to 0.007) |
| PRS * Age * Scale |  | -0.003 (-0.01 to -0.0005) |
| PRS * Scale * Reporter |  | 0.03 (-0.01 to 0.06) |
| PRS * Reporter * Age |  | -0.2 (-0.02 to -0.01) |
| Reporter * Sex |  | -0.12 (-0.17 to -0.07) |
| Reporter * Scale |  | -0.24 (-0.28 to -0.19) |
| Reporter * Scale * Sex |  | -0.17 (-0.24 to -0.11) |
| Reporter * age |  | 0.03 (0.02 to 0.04) |
| Sex * Age |  | 0.01 (0.01 to 0.02) |
| Reporter * Sex * Age |  | -0.04 (-0.05 to -0.02) |
| Scale * Age |  | -0.02 (-0.025 to -0.020) |
| Scale * Sex |  | -0.20 (-0.23 to -0.16) |
| Scale * Age * Sex |  | -0.03 (-0.03 to -0.02) |
| Maternal education | *1=low education, 2=average education, 3=high education* | -0.04 (-0.05 to -0.02) |
| Maternal social class | *0=low, 1=high* | 0.04 (0.01 to 0.06) |
| Mother's age |  | -0.01 (-0.012 to -0.01) |
| First PC |  | 0.47 (-0.62 to 1.57) |
| Second PC |  | -2.06 (-3.82 to -0.29) |

|  |  |
| --- | --- |
| Third PC | 0.41 (-0.65 to 1.46) |
| Fourth PC | 0.18 (-0.88 to 1.23) |
| Fifth PC | -0.37 (-1.41 to 0.66) |
| Intercept | 1.38 (1.30 to 1.47) |
| **Random-effects Parameters** | **Variance components (95% CIs)** |
| **Unstructured individual child level** |  |
| Sd (age_8y) | 0.05 (0.04 to 0.05) |
| Sd (_cons) | 0.32 (0.31 to 0.33) |
| Corr (age_8y, cons) | 0.57 (0.51 to 0.62) |
| **Unstructured scale-level** |  |
| Sd (reporter) | 0.56 (0.55 to 0.58) |
| Sd (_cons) | 0.38 (0.37 to 0.39) |
| Corr (reporter, cons) | -0.47 (-0.50 to -0.44) |
| Sd (Residual) | 0.46 (0.45 to 0.46) |
| AIC<sup>a</sup> | 102130.7 |
| Log-likelihood | -51029.34 |

**Legend:** <sup>a</sup>Akaike Information Criterion. Reporter: 0 indicates parents and 1 indicates teachers; sex: 0 indicates boys and 1 indicates girls; scale: 0 indicates internalising and 1 indicates externalising; maternal education: 1 indicates low education (equivalent to O level, Certificate of Secondary Education (CSE) or vocational training), 2 indicates A level, and 3 indicates some degree; Mother's age: Maternal age in years during pregnancy.
†Outcome is transformed into the log-scale and adding 1: $\ln(x+1)$ . The coefficients (apart from the intercept) represent the percentage change in the outcome. By exponentiating the intercept and removing 1 ( $\exp(x)-1$ ), the intercept will indicate the mean score on the actual internalising subscale at 8 years of age in boys and when reported by the parent.
Mean differences on the actual scale scores between low and high PRS at the youngest and oldest ages are reported in **Supplementary Table 15**.

**Figure 2.**
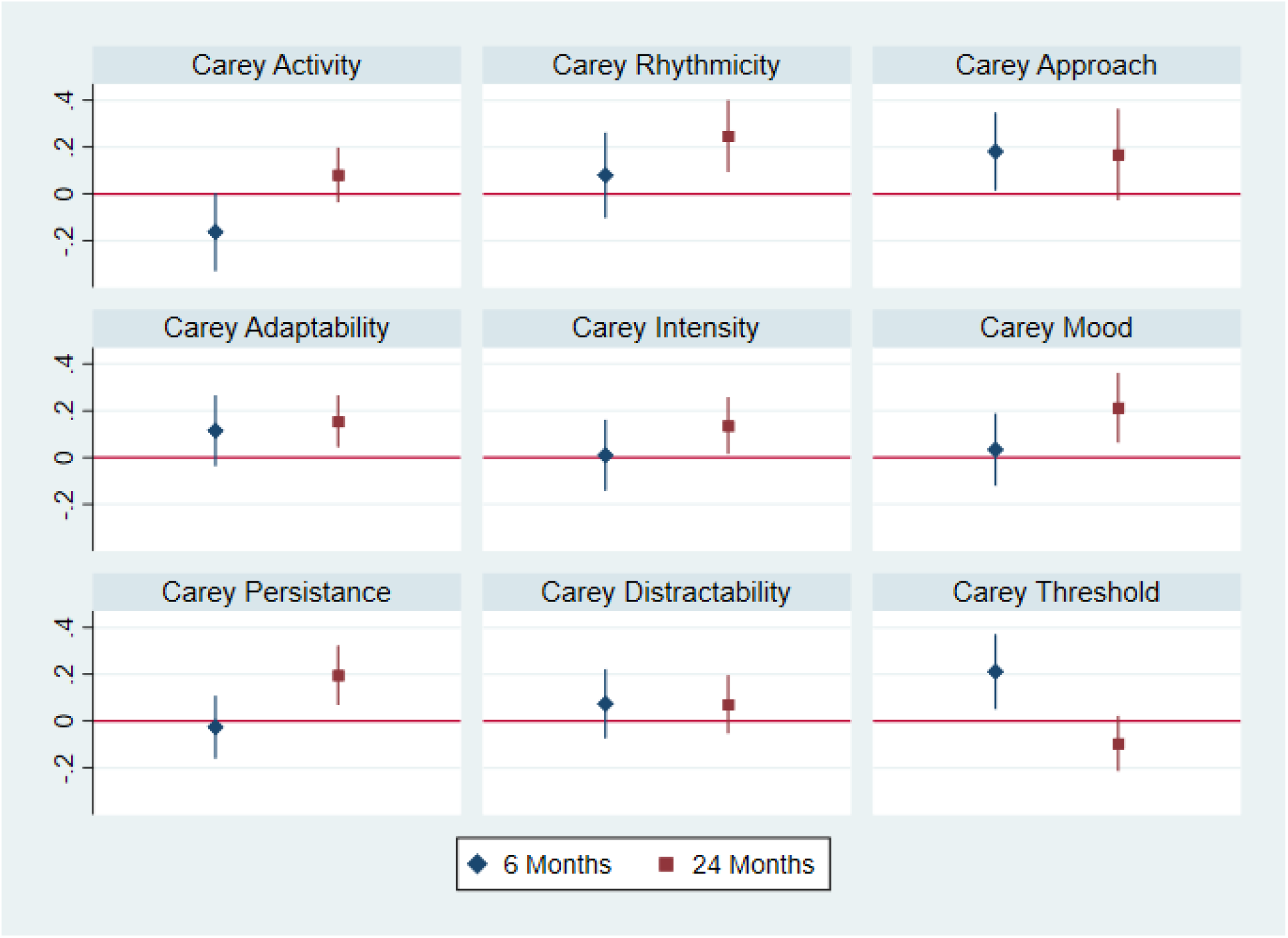
Graphical representation of separate linear regressions of child PRS on CTS at 6 and 24 months. All effect estimates were adjusted for child’s sex and first five principal components of genetic ancestry. The outcome scales were already adjusted for gestational age and child’s age. 95% CIs were not adjusted for multiple testing, therefore the evidence for some of these associations may be weaker than illustrated in the figure.

**Figure 3.**
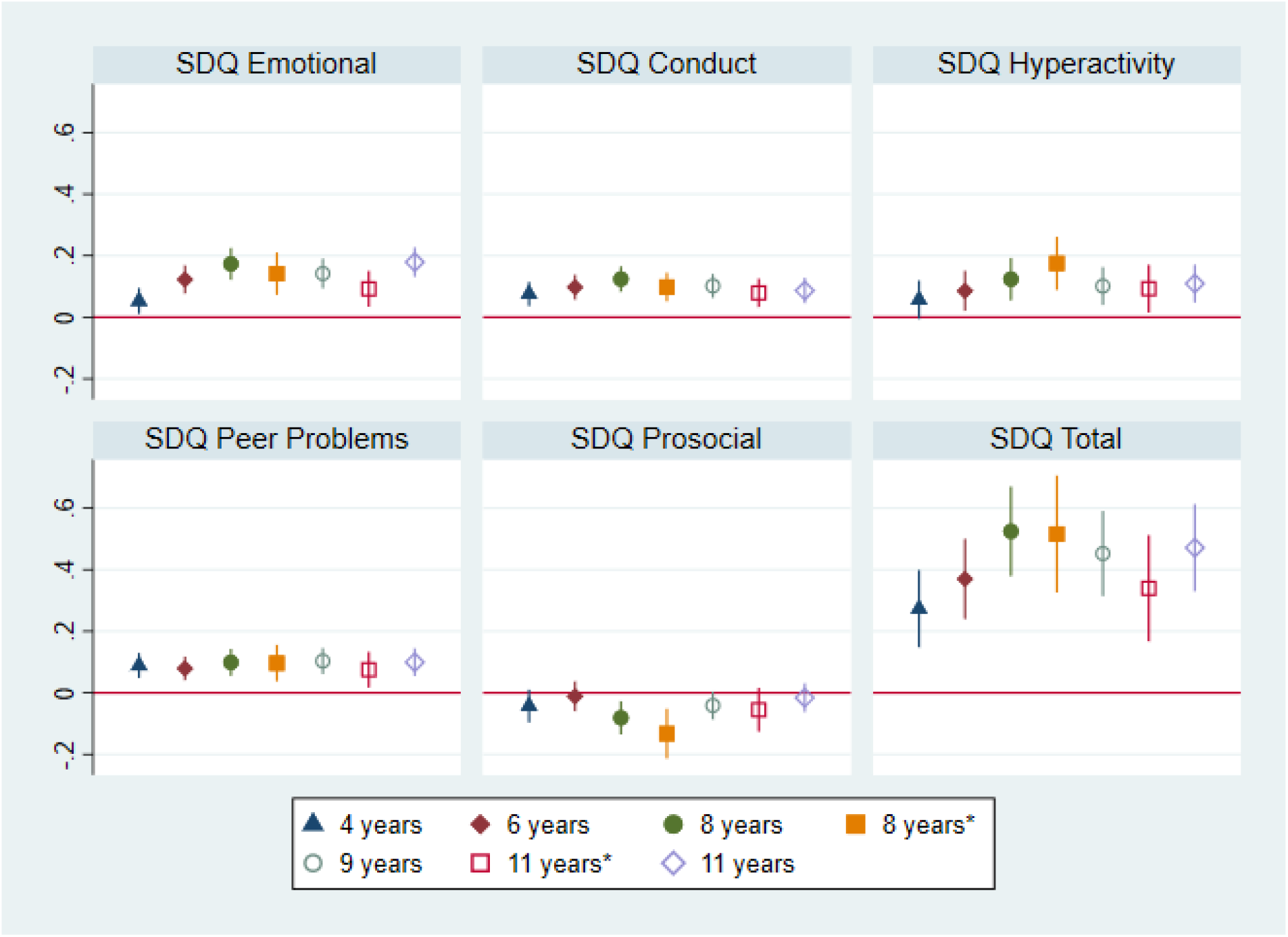
Graphical representation of separate linear regressions of child PRS on SDQ at different ages. All effect estimates were adjusted for child’s age, sex and first five principal components of genetic ancestry. Estimates obtained signalled with an asterisk* were obtained by SDQ assessed by the teacher. 95% CIs were not adjusted for multiple testing, therefore the evidence for some of these associations may be weaker than illustrated in the figure.

**Figure 4.**
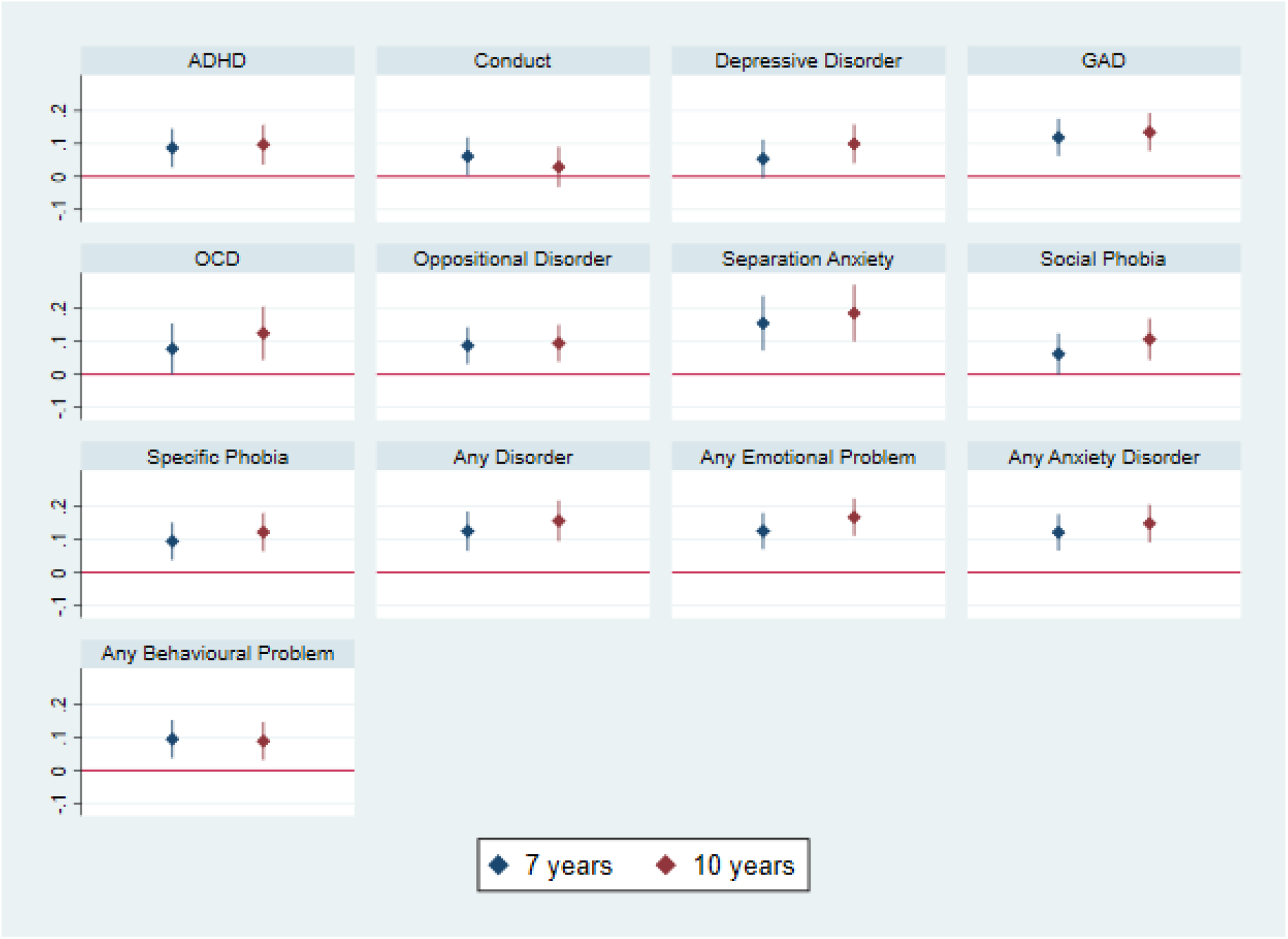
Graphical representation of separate ordinal regressions child PRS on DAWBA subscales at 7 and 10 years of age. An increase in the log odds indicates an increased risk of being in a higher DAWBA category. 95% CIs were not adjusted for multiple testing, therefore the evidence for some of these associations may be weaker than illustrated in the figure.

**Figure 5.**
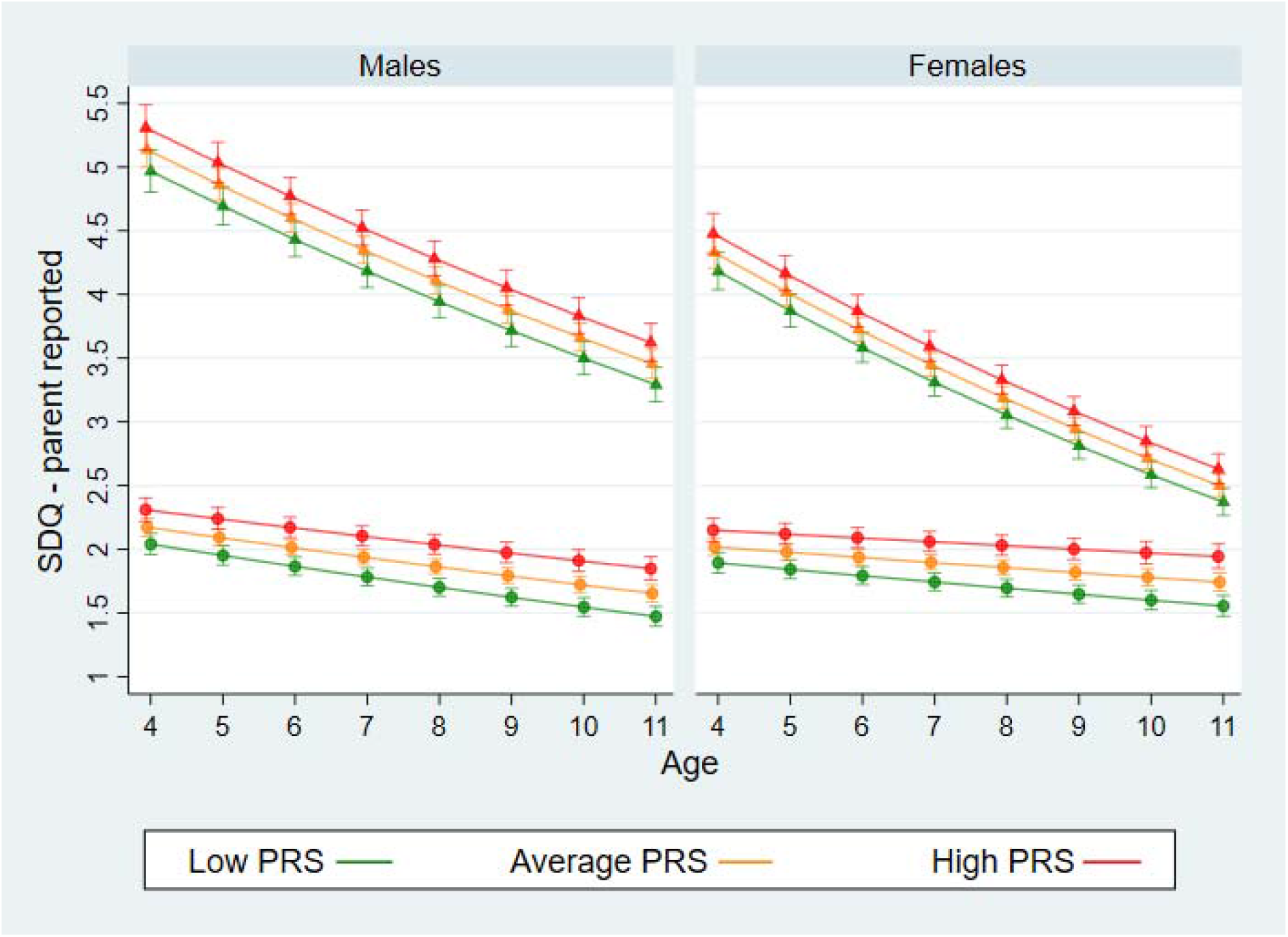
Trajectories of internalising and externalising problems by PRS levels across age when SDQ is mother reported by sex of the child. Low PRS corresponds to 1SD lower than average score, Average PRS corresponds to the mean PRS score, and high PRS corresponds to 1 SD higher than average. Circles represent the internalising scale and the triangle the externalising.

**Figure 6.**
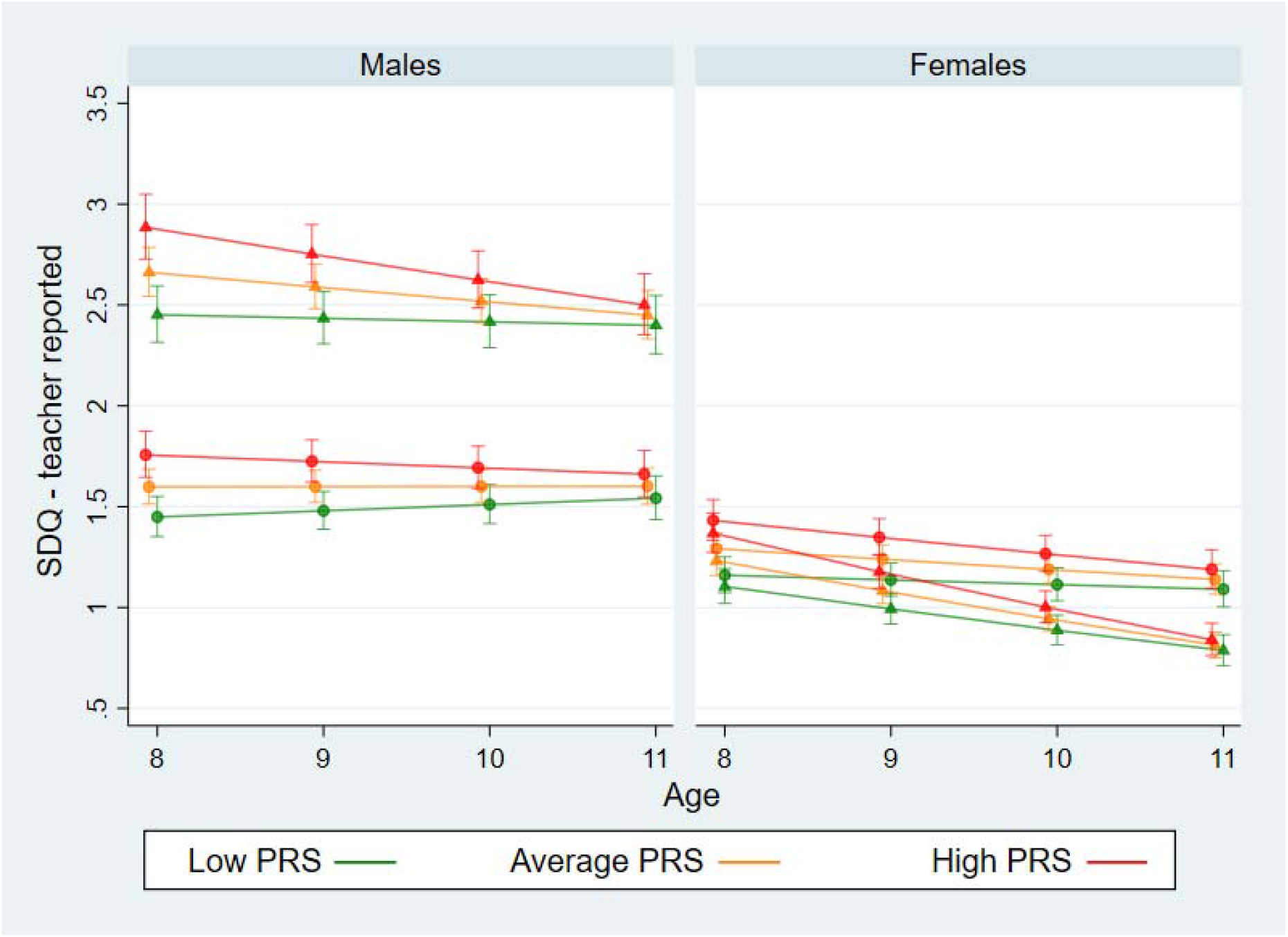
Trajectories of internalising and externalising problems by PRS levels across age when SDQ is teacher reported by sex of the child. Low PRS corresponds to 1SD lower than average score, Average PRS corresponds to the mean PRS score, and high PRS corresponds to 1 SD higher than average. Circles represent the internalising scale and the triangle the externalising.

### Sensitivity analyses

In this study we hypothesised that there could be differential misclassification of child outcomes dependent on maternal PRS for neuroticism.

First, we found little evidence of an association between maternal PRS for neuroticism and child’s behavioural and emotional problems as rated by the teacher, as assessed by 95% confidence intervals overlapping with the null (**Supplementary Table 8**). The maternal PRS for neuroticism had little or no influence on teachers’ assessment of child’s behaviours once accounting for the child’s own PRS, minimising the likelihood that mothers with a higher PRS for neuroticism may have influenced the teacher’s opinion of the child (e.g., through their behaviours during scholastic meetings). Second, while we found that mothers and teachers did indeed rate child’s emotional and behavioural problems differently (mean of the difference in score ≠ 0) (**Supplementary Table 9**), with mothers rating the child’s behavioural and emotional problems higher than teachers, when we examined whether maternal PRS was associated with a “difference score”, we did not find any convincing evidence to support this hypothesis. In addition, effect estimates were small and with differing directions of effect, suggesting that the association of the maternal PRS with “difference in score” may be negligible in our sample (**Supplementary Table 10**).

To investigate whether our findings were compatible with a causal association of the child PRS with both emotional and behavioural problems we performed two sensitivity analyses. First, to estimate a potential causal association between the child PRS and later emotional and behavioural problems we adjusted for the maternal PRS for neuroticism (i.e., a potential confounder of this association). Our effect estimates remained consistent across most of the results (**Supplementary Table 11)**. However, incorporating the maternal PRS into the model led to larger confidence intervals around our effect estimates, reducing their precision.

Second, to estimate a potential causal association between the child PRS for neuroticism and later emotional and behavioural problems we re-performed our single-level analyses using a PRS constructed using a genome-wide significant P-value threshold with and without adjustment for the maternal PRS for neuroticism (**Supplementary Table 12**). Effect estimates decreased across all our findings, particularly for those measured at earlier ages. Very weak evidence was found at 4 years of age for an association between the child PRS for neuroticism and the total scores of emotional and behavioural difficulties (β=0.05, 95% CI: -0.14 to 0.24). However, as early as at 6 years, we found some evidence of an association between the child neuroticism PRS and the total scores of emotional and behavioural difficulties (β=0.22, 95% CI: 0.02 to 0.42). We also found evidence for an association between the child PRS for neuroticism and a higher risk of separation anxiety, both when assessed using the DSM-IV and ICD-10 criteria (OR: 1.21, 95% CI: 1.05 to 1.37; OR: 1.39, 95% CI: 1.17 to 1.62), social phobia (OR: 1.17, 95% CI: 1.05 to 1.29) and specific phobia (OR: 1.16, 95% CI: 1.05 to 1.27). These results remained largely consistent upon adjusting for the maternal PRS for neuroticism.

### Missing data

**Results** obtained in the 100 multiply imputed datasets are presented in the **Supplementary Materials** (see **Supplementary Table 13** for auxiliary variables and **Supplementary Table 14** for findings generated from imputed datasets). Monte Carlo errors were <10% of the standard error and fraction of missing information (FMI) values were no larger than 11 in the Carey models imputation and 54 in the SDQ model imputation, both indicating an adequate level of statistical reproducibility of the multiply imputed analyses ^47^. Findings obtained in the multiply imputed datasets were largely consistent (with the most different result observed for the total score of the SDQ at 9 years: N complete case analysis (cca)= 5,516, β_cca_: 0.45, 95% CI_cca_: (0.31 to 0.59), P-value_cca_=2.98 × 10-10, N imputed (i)= 7,451, β_i_: 0.50, 95% CI_i_: (0.37 to 0.63), P-value_i_=4.76 × 10-14) with those reported in the CCA, suggesting that the levels of attrition did not substantially bias our effect estimates under the assumption that data were MAR.

## Discussion

Psychiatric genetic studies have recently started investigating associations between genetic variants linked to psychological and psychiatric outcomes in adults on their correlates in children and adolescents ^17,48^. To the best of our knowledge, this is the first study to have examined how a PRS for neuroticism is manifested phenotypically throughout early infancy to late childhood in addition to the first to examine its association with trajectories of internalising and externalising symptoms across these age groups.

In this study, we found evidence of an association between a higher child PRS for neuroticism and a host of behavioural and emotional problems across different ages and with some suggestion of an association appearing as early as 6 months of age, though evidence supporting this association weakened once considering multiple testing. In addition, when estimating trajectories of child internalising and externalising problems between 4 and 11 years of age, we found that they both reduced with age, in line with early childhood emotional issues often regulating by late childhood. Both girls and boys had higher overall levels of externalising, as compared to internalising, problems; however, the reduction of externalising trajectories over time was sharper than for internalising trajectories. Furthermore, we found strong evidence that a higher PRS for neuroticism was associated with an increase in the overall level of internalising problems but that the magnitude of effect was halved for the externalising scale. We also found evidence that a higher PRS for neuroticism was associated with smaller rates of reduction of internalising problems, suggesting a dampened recovery from the emotional problems experienced in children with higher levels of this PRS (**Figure 5**). In addition, we found evidence that teachers gave lower scores than parents on internalising and externalising scales to both boys and girls. However, the rate of decrease was also influenced by the reporter of the scale, with the teacher reporting smaller rates of reduction compared to parents (**Figure 6**). Lastly, we found evidence that both parents and teachers scored girls lower on externalising problems and higher on internalising problems as compared to boys. These findings are consistent with psychological and psychiatric literature which has identified higher levels of total difficulties across childhood in boys as compared to girls ^49,50^ and literature on teachers reporting fewer problems than parents ^49,51^.

Our findings suggest that a child PRS for neuroticism has larger effects as the child ages. Here, we hypothesise two plausible explanations for this observation: first, the child PRS for neuroticism was derived using a GWAS which captures neuroticism as measured in adulthood and not in childhood and thus, as the child becomes older, the phenotype continues to approach the measured construct. This means that the emotional and behavioural measured constructs may have suffered from higher measurement error in earlier ages (e.g., because behavioural expression of emotional problems is more undifferentiated) than when measured later in childhood. Secondly, the child PRS for neuroticism may be increasingly expressed with age in line with previous genetically informed longitudinal studies (such using a twin-design) ^52^ that have found that genetic contribution to personality traits such as neuroticism and extraversion reaches its peak at around 40 years of age (see Kandler ^53^ for a review of these studies).

Across scales, the largest effect sizes were found for internalising problems and anxiety-related clinical disorders. Nonetheless, our analyses also found evidence that the child neuroticism PRS was associated with increased behavioural or externalising problems (e.g., conduct problems or hyperactivity).

Whereas the findings for the internalising problems were largely expected because anxiety and low mood represent different facets of neuroticism, the strong evidence of an association with externalising problems, particularly at younger ages, highlights the importance of considering behavioural problems as a manifestation of a potential underlying emotional disturbance. These results align with a variety of research supporting a substantial overlap between neuroticism and both internalising and externalising psychopathology ^7,54,55^. This may be particularly important in the context of interventions where the externalising/behavioural problems are the target of the intervention ^56^. Specifically, for those interventions where little attention is given to potential underlying anxiety and depressive disorders which may be sustaining the behavioural problems and may still persist once those are reduced. In fact, a PRS for neuroticism may be expressed in different ways during childhood as externalising symptoms may be the first reaction of an underlying internalising problem, which could potentially reflect two phenotypes representing distinct traits or simply representing different modalities in which an underlying common factor (e.g., neuroticism) is expressed.

Finally, we found strong evidence of an association between the child PRS for neuroticism and other psychological traits (e.g., locus of control and self-esteem) and IQ. As IQ and self-esteem were measured at the same age, it is difficult to speculate whether this observed association between the PRS for neuroticism and IQ could be partially mediated through low scholastic self-esteem or whether this association reflects an effect in the reverse direction (i.e., an association between the PRS and low scholastic self-esteem is mediated by performance on IQ tests).

When testing whether our results were compatible with causality (i.e., the effect of the child PRS for neuroticism on our outcomes of interest) by adjusting for maternal PRS (i.e., a potential confounder of these associations), the strength of the evidence attenuated across all findings but effects sizes remained comparable with the unadjusted complete case analyses. In addition, findings for an association between the child PRS and emotional and anxiety related disorders remained robust to this adjustment. Conversely, when we re-constructed the neuroticism PRS using a genome-wide significance threshold to explore whether findings may be more likely to be driven through genetic liability to neuroticism *per se*, we found sharp reductions in all our effect estimates, with little evidence of association remaining for the emotional problems as identified by the SDQ and the anxiety-specific related problems as identified by the DAWBA. These findings suggest that our results are unlikely to be entirely mediated via genetic liability to neuroticism but likely reflect the effect of pleiotropic pathways from the neuroticism PRS to outcomes examined.

## Strengths and limitations

This study has several strengths. First, we used a large, well-characterised, prospective population-based sample to examine neuroticism-related phenotypes during childhood, a key period of development that closely predates the start of the peak in incidence of internalising problems (i.e., early adolescence)^57^. Second, we employed multiple measures of psychological problems at different ages during childhood and we compared the effect of different reporters on the scores to account for possible misclassification of the outcomes. Third, when trying to estimate the potential causal association between the child PRS and various psychological outcomes, maternal and paternal PRS for neuroticism represent potentially important confounding factors as both scores will correlate with the child’s PRS which could have influenced manifestation of emotional and behavioural problems in the child via direct (e.g., genetic effect) and indirect (e.g., mental health phenotype, parenting, broader familiar environment) pathways. Here, as recommended in Tubbs et al ^58^, when trying to estimate the causal association between the child PRS and an outcome that could be confounded by parental genetics, we adjusted for at least one of the available PRSs (in our case, the maternal PRS for neuroticism). However, it is important to emphasise that if paternal genotype confounds this association, the child estimate will be biased toward the paternal effect, albeit to a lesser extent that if maternal genotype were not adjusted for. Fourth, our results remained robust to a variety of sensitivity analyses testing attrition and potential differential misclassification.

Our study also has several potential limitations. First, there were moderate to high levels of missingness, which may have led to reduced statistical power to detect small effects in addition to potentially introducing bias into our analyses. However, the findings obtained in the imputed datasets were comparable to those obtained using a complete case analysis, indicating that any bias present due to missingness was likely to be minimal, under the assumption that data were missing-at-random. Second, children’s emotional and behavioural problems were mostly reported by the parent (i.e., primarily the mother), especially at earlier ages of the child when the teacher rating was not available. As the mother shares genetics with the child, we hypothesised that differential misclassification of the outcome depending on maternal PRS could have occurred (**Supplementary Figure 1**). In fact, previous studies support the existence of a “distortion model” ^59–61^, for which reporter’s mental health characteristics may be systematically biasing the rating of child emotional and behavioural problems. However, after conducting a variety of sensitivity analyses, we generally found little evidence that misclassification according to the maternal PRS to neuroticism was present. In fact, conflicting research suggests that the magnitude of these biases is often negligible ^23^; in addition, some of this work indicates that differences in behaviours linked to reporters may be the result of mechanisms other than misclassification. Here, we speculate on a few potential mechanisms. First, differences in scores due to reporters (mother vs teacher) may reflect actual differences in behaviour of children across different situational contexts (i.e., cross-situational differences) ^62^, where the child manifests certain behaviours and emotional reactions with higher frequency and/or intensity in a familiar context than in a structured environment such as school. Second, differences in scores may relate to different abilities of parents and teachers in recognising psychological problems of children. In fact, parents may be more sensitive to small fluctuations in mood than teachers who need to pay attention to multiple children simultaneously. Third, parents and children share their genetics; this could open other pathways through which child’s emotional and behavioural problems are manifested differently when in the presence of the parent. In fact, parents may be evoking different behavioural and emotional responses in children both via direct (i.e., genetic confounding) and indirect (e.g., parenting practices) genetic effects^63^. For example, a mother or a father with a high PRS for neuroticism may use more discipline and/or harsher parenting practices which may, in turn, evoke more conduct problems or more withdrawal reactions in the child. In this scenario, a parental PRS would be acting as a confounding factor and parenting practices would be on the causal pathway as mediators of the association (**Supplementary Figure 3**). Dysfunctional parenting practices could reflect both underlying mental health problems (e.g., anxiety and depression) and/or a direct response to a child’s behaviour. Similarly, a child PRS as expressed via temperamental difficulties and/or more dysfunctional behaviours may have an evocative effect on parents, eliciting worse parenting practices or poorer mental health in the parents, which could reinforce a dysfunctional cycle. Nevertheless, exploring evocative genetic effects is beyond the scope of this study. Another potential limitation of this study regards its generalisability. Whereas replicating this study in another longitudinal cohort with similar demographics may provide more robustness to the generalisability of these findings to WEIRD (Western, Educated, Industrialised, Rich and Democratic)^64^ populations, the relevance of findings from this study to the full range of human variation and diversity is unknown ^65^. Finally, although increases in the sample size of GWASs have led to increased statistical power, the overall variance explained by SNPs used to develop PRSs for neuroticism remains small (around 3%)^66^, limiting the ability of such PRSs to identify small to modest effects of genetic liability to neuroticism on subsequent emotional and behavioural outcomes.

## Conclusions

Our comprehensive analyses using a large, well-characterised birth cohort study of up to 6,271 children suggest that it is possible to detect manifestations of a PRS for neuroticism on temperamental traits, internalising and externalising problems and a variety of clinical disorders as early as in infancy and childhood. Furthermore, we found that a child PRS for neuroticism was associated with trajectories of internalising and externalising problems, influencing both their overall levels and their rate of change across time. These findings, if successfully replicated in independent samples, could be used to inform development of prediction models for later-life psychopathology in high-risk groups. In addition, future studies with complete parental genotype and different assessors of those psychological outcomes evaluated in this study may help to further elucidate the causal nature of the associations reported. Finally, the use of trio designs combined with phenotypic measures of early-life environmental factors may help to further disentangle the direct and indirect pathways leading from parental genotype to negative psychological outcomes in childhood.

## Data Availability

The data analysed in this study is subject to the following licenses/restrictions: ALSPAC datasets can be accessed pending approval of research proposals. Requests to access these datasets should be directed to.

## Ethics Statement

The studies involving human participants were reviewed and approved by Ethical approval for the study was obtained from the ALSPAC Law and Ethics Committee and Southwest National Health Service (NHS) Research Ethics Committee; participants gave written informed data consent. Written informed consent to participate in this study was provided by the participants’ legal guardian/next of kin.

## Author Contributions

IC contributed to conception of the study. IC, DK, KT and RP contributed to the design of the study. HS derived the PRS at all its thresholds. IC performed the statistical analyses and wrote the first draft of the manuscript. All authors contributed to manuscript revision, read, and approved the submitted version.

## Acknowledgments

We are extremely grateful to all the families who took part in this study, the midwives for their help in recruiting them, and the whole ALSPAC team, which includes interviewers, computer and laboratory technicians, clerical workers, research scientists, volunteers, managers, receptionists, and nurses.

