## Supplementary Materials for "Childhood trajectories of internalising and externalising problems associated with a polygenic risk score for neuroticism in a UK birth cohort study"

[Supplementary Table 12. Associations between child PRS for neuroticism (threshold P < 5 × 10^−8^) and all psychological outcomes unadjusted and adjusted for maternal PRS for neuroticism.	39](#_Toc90486467)

### Supplementary information on genotyping

### Genotype Information

ALSPAC children were genotyped using the Illumina HumanHap550 quad chip genotyping platforms. The resulting raw genome-wide data were subjected to standard quality control methods. Individuals were excluded on the basis of gender mismatches; minimal or excessive heterozygosity; disproportionate levels of individual missingness (>3%) and insufficient sample replication (IBD < 0.8). Population stratification was assessed by multidimensional scaling analysis and compared with Hapmap II (release 22) European descent (CEU), Han Chinese, Japanese and Yoruba reference populations; all individuals with non-European ancestry were removed. SNPs with a minor allele frequency of < 1%, a call rate of < 95% or evidence for violations of Hardy-Weinberg equilibrium (P < 5 x 10^-7^) were removed. Cryptic relatedness was measured as proportion of identity by descent (IBD > 0.1). Related subjects that passed all other quality control thresholds were retained during subsequent phasing and imputation. 9,115 subjects and 500,527 SNPs passed these quality control filters.

ALSPAC mothers were genotyped using the Illumina human660W-quad array at Centre National de Génotypage (CNG) and genotypes were called with Illumina GenomeStudio. PLINK (v1.07) was used to carry out quality control measures on an initial set of 10,015 subjects and 557,124 directly genotyped SNPs. SNPs were removed if they displayed more than 5% missingness or a Hardy-Weinberg equilibrium P value of less than 1.0 x 10^-6^. Additionally, SNPs with a minor allele frequency of less than 1% were removed. Samples were excluded if they displayed more than 5% missingness, had indeterminate X chromosome heterozygosity or extreme autosomal heterozygosity. Samples showing evidence of population stratification were identified by multidimensional scaling of genome-wide identity by state pairwise distances using the four HapMap populations as a reference, and then excluded. Cryptic relatedness was assessed using a IBD estimate of more than 0.125 which is expected to correspond to roughly 12.5% alleles shared IBD or a relatedness at the first cousin level. Related subjects that passed all other quality control thresholds were retained during subsequent phasing and imputation. 9,048 subjects and 526,688 SNPs passed these quality control filters.

After combining genotype data in the mothers and the children, SNPs with genotype missingness above 1% were removed due to poor quality (11,396 SNPs removed) and a further 321 subjects were removed due to potential ID mismatches. This resulted in a dataset of 17,842 subjects. Imputation of the target data was performed using Impute V2.2.2 against the 1000 genomes reference panel (Phase 1, Version 3) (all polymorphic SNPs excluding singletons), using all 2186 reference haplotypes (including non-Europeans). This gave 8,237 eligible children and 8,196 eligible mothers with available genotype data after exclusion of related subjects using cryptic relatedness measures described previously.

### Supplementary Table 1. Number of SNPs included in the analyses at different P-value thresholds.

| **P-value threshold** | **Child PRS** | **Maternal PRS** |
| --- | --- | --- |
| **5.00 x 10^-8^** | **113** | **114** |
| 1.00 x 10^-7^ | 140 | 141 |
| 1.00 x 10^-6^ | 235 | 236 |
| 1.00 x 10^-5^ | 492 | 493 |
| 0.0001 | 1226 | 1225 |
| 0.001 | 3771 | 3767 |
| 0.01 | 14439 | 14424 |
| **0.05** | **39401** | **39352** |
| 0.1 | 60857 | 60797 |
| 0.2 | 93822 | 93839 |
| 0.3 | 119770 | 119755 |
| 0.4 | 141472 | 141409 |
| 0.5 | 159446 | 159269 |
| 1 | 211672 | 211648 |

**Legend:** We have highlighted in bold the two P-value thresholds employed in this work.

### Supplementary information on outcome measures

#### Carey Infant Temperament Scales (CTSs) – *6 and 24 months*

The analyses were performed separately across every subscale relating to the temperament domains previously identified ^1^: Activity (i.e., motor component in a child’s functioning), Rhythmicity (i.e., regularity; predictability in daily functions), Approach (i.e., initial response to a novel stimulus), Adaptability (i.e., behavioural flexibility in changing environments), Intensity (i.e., energy level of an emotional response), Mood (i.e., tone of overall affect: positive or negative), Persistence (i.e., continuation of activity in face of obstacles), Distractibility (i.e., effectiveness of extraneous stimuli in altering the direction of ongoing attention) and Threshold (i.e., the intensity level of stimulation that is necessary to evoke a discernible response) ^2^. The measure was predominantly mother reported (94%) at 6 and 24 months of age. In the current sample, internal reliability as measured with Cronbach alpha was mixed, ranging from poor to good (0.36 to 0.73) when measured at 6 months and was more satisfactory when measured at 24 months (0.59 to 0.87).

#### Strengths and Difficulties Questionnaire (SDQ) – *4 to 11 years*

The total score used here is a weighted sum of the components of the Hyperactivity, Emotional Symptoms, Conduct Problems and Peer Problems scores. This questionnaire was used to assess child difficulties at 4 years (parent report – 98% mother report), 6 years (parent report – 97% mother report), 8 years (both parent – 97.8% mother report – and teacher report), 9 years (parent report – 97.6% mother report -) and at 11 years of age (both parent – 97.6% mother report – and teacher report). Internalising and externalising subscales were derived summing up the scores from the Emotional Symptoms and Peer Problems and the Hyperactivity and Conduct Problems subscales, respectively. Subscale scores were pro-rated if 1 or 2 items were missing ([www.sdqinfo.com/b4.html](http://www.sdqinfo.com/b4.html)). In the current sample, Cronbach’s alpha values for the total SDQ score were 0.80, 0.79, 0.82, 0.81, and 0.81, at age 4, 6, 8, 9, and 11 years, respectively when reported by the parent and 0.87 and 0.88 at age 8 and 11 years, respectively, when reported by the teacher.

#### Axis-I Disorders – *7 and 11 years*

The DAWBA has incorporated information from both parent and teacher reports for the ADHD and oppositional/conduct disorders, which should have minimised the possibility of differential misclassification. The advantages of using the ordered categorical variables over a binary classification of the scale are described elsewhere ^3^.

### Supplementary Table 2. Descriptives of outcomes measures.

| **Variable** | **Mean** | **SD** | **Min** | **Max** | **Mean (SD) age at completion in months** | **Cronbach’s alphas** | **Reporter** | **All Participants with Measure** | **Genotyped Participants** |
| --- | --- | --- | --- | --- | --- | --- | --- | --- | --- |
| Sex |  |  | 0 | 1 |  |  |  | 15,039 (48.86% females) | 7,847 (48.74% females) |
| Carey Activity | 40.13 | 6.34 | 13 | 60 | 6.09 (0.31) | 0.56 | Mother | 10,503 | 6,275 |
| Carey Rhythm | 15.97 | 6.93 | 0 | 46 | 6.09 (0.31) | 0.73 | Mother | 10,500 | 6,273 |
| Carey Approach | 15.01 | 6.36 | 0 | 43 | 6.09 (0.31) | 0.71 | Mother | 10,495 | 6,270 |
| Carey Adapt | 14.00 | 5.71 | 0 | 42 | 6.09 (0.31) | 0.59 | Mother | 10,508 | 6,275 |
| Carey Intensity | 25.03 | 5.59 | 4 | 49 | 6.09 (0.31) | 0.36 | Mother | 10,494 | 6,273 |
| Carey Mood | 15.81 | 5.91 | 0 | 44 | 6.09 (0.31) | 0.61 | Mother | 10,496 | 6,271 |
| Carey Persist | 13.57 | 5.11 | 0 | 34 | 6.09 (0.31) | 0.60 | Mother | 10,504 | 6,273 |
| Carey Distract | 14.02 | 5.61 | 0 | 36 | 6.09 (0.31) | 0.58 | Mother | 10,506 | 6,275 |
| Carey Threshold | 27.51 | 6.06 | 3 | 50 | 6.09 (0.31) | 0.49 | Mother | 10,490 | 6,268 |
| Carey Activity | 23.15 | 4.54 | 3 | 36 | 24.44 (1.13) | 0.59 | Mother | 10,327 | 6,264 |
| Carey Rhythm | 12.79 | 5.76 | 0 | 40 | 24.44 (1.13) | 0.80 | Mother | 10,321 | 6,261 |
| Carey Approach | 20.23 | 7.68 | 0 | 44 | 24.44 (1.13) | 0.83 | Mother | 10,318 | 6,257 |
| Carey Adapt | 12.79 | 4.18 | 0 | 28 | 24.44 (1.13) | 0.75 | Mother | 10,272 | 6,237 |
| Carey Intensity | 21.40 | 4.54 | 4 | 36 | 24.44 (1.13) | 0.81 | Mother | 10,317 | 6,259 |
| Carey Mood | 18.08 | 5.69 | 1 | 48 | 24.44 (1.13) | 0.83 | Mother | 10,325 | 6,263 |
| Carey Persist | 16.26 | 4.89 | 0 | 35 | 24.44 (1.13) | 0.82 | Mother | 10,312 | 6,256 |
| Carey Distract | 24.54 | 4.68 | 0 | 40 | 24.44 (1.13) | 0.87 | Mother | 10,319 | 6,259 |
| Carey Threshold | 19.05 | 4.40 | 0 | 32 | 24.44 (1.13) | 0.72 | Mother | 10,327 | 6,264 |
| IQ (WISC) | 103.97 | 16.54 | 45 | 151 | 103.83 (3.92) | - | Child | 7,354 | 5,297 |
| Locus of control | 5.99 | 2.08 | 0 | 12 | 103.83 (3.92) | 0.47 | Child | 6,380 | 4,601 |
| **Variable** | **Median** | **IQR** | **Min** | **Max** | **Mean (SD) age at completion** | **Alpha** | **Reporter** | **All Participants with Measure** | **Genotyped Participants** |
| SDQ total score | 8 | 6 - 12 | 0 | 33 | 48 (1.44) | 0.80 | Mother | 9,457 | 5,911 |
| SDQ total score | 7 | 4 - 10 | 0 | 33 | 81.45 (1.36) | 0.79 | Mother | 8,404 | 5,514 |
| SDQ total score | 7 | 4 - 11 | 0 | 38 | 98.43 (3.09) | 0.82 | Mother | 7,785 | 5,287 |
| SDQ total score | 4 | 2 - 9 | 0 | 37 | 99.92 (3.74) | 0.87 | Teacher | 6,368 | 3,589 |
| SDQ total score | 6 | 3 - 9 | 0 | 35 | 115.82 (1.57) | 0.81 | Mother | 8,040 | 5,520 |
| SDQ total score | 4 | 1 - 9 | 0 | 37 | 133.99 (3.89) | 0.88 | Teacher | 7,666 | 4,273 |
| SDQ total score | 5 | 3 - 9 | 0 | 34 | 140.62 (1.64) | 0.81 | Mother | 7,354 | 5,131 |
| SDQ emotional subscale | 1 | 0 - 2 | 0 | 10 | 48 (1.44) | 0.68 | Mother | 9,500 | 5,932 |
| SDQ emotional subscale | 1 | 0 - 2 | 0 | 10 | 81.45 (1.36) | 0.63 | Mother | 8,423 | 5,521 |
| SDQ emotional subscale | 1 | 0 - 3 | 0 | 10 | 98.43 (3.09) | 0.69 | Mother | 7,795 | 5,286 |
| SDQ emotional subscale | 1 | 0 - 2 | 0 | 10 | 99.92 (3.74) | 0.80 | Teacher | 6,369 | 3,590 |
| SDQ emotional subscale | 1 | 0 - 2 | 0 | 10 | 115.82 (1.57) | 0.68 | Mother | 8,052 | 5,528 |
| SDQ emotional subscale | 0 | 0 - 2 | 0 | 10 | 133.99 (3.89) | 0.80 | Teacher | 7,665 | 4,273 |
| SDQ emotional subscale | 1 | 0 - 2 | 0 | 10 | 140.62 (1.64) | 0.67 | Mother | 7,340 | 5,121 |
| SDQ hyperactivity subscale | 4 | 2 - 5 | 0 | 10 | 48 (1.44) | 0.76 | Mother | 9,492 | 5,931 |
| SDQ hyperactivity subscale | 3 | 1 - 5 | 0 | 10 | 81.45 (1.36) | 0.77 | Mother | 8,409 | 5,522 |
| SDQ hyperactivity subscale | 3 | 1 - 5 | 0 | 10 | 98.43 (3.09) | 0.80 | Mother | 7,796 | 5,286 |
| SDQ hyperactivity subscale | 2 | 0 - 4 | 0 | 10 | 99.92 (3.74) | 0.87 | Teacher | 6,369 | 3,590 |
| SDQ hyperactivity subscale | 3 | 1 - 4 | 0 | 10 | 115.82 (1.57) | 0.76 | Mother | 8,069 | 5,542 |
| SDQ hyperactivity subscale | 1.5 | 0 - 4 | 0 | 10 | 133.99 (3.89) | 0.88 | Teacher | 7,666 | 4,273 |
| SDQ hyperactivity subscale | 2 | 1 - 4 | 0 | 10 | 140.62 (1.64) | 0.77 | Mother | 7,339 | 5,121 |
| SDQ conduct subscale | 2 | 1 - 3 | 0 | 10 | 48 (1.44) | 0.58 | Mother | 9,486 | 5,928 |
| SDQ conduct subscale | 1 | 0 - 2 | 0 | 10 | 81.45 (1.36) | 0.55 | Mother | 8,433 | 5,529 |
| SDQ conduct subscale | 1 | 0 - 2 | 0 | 10 | 98.43 (3.09) | 0.59 | Mother | 7,798 | 5,287 |
| SDQ conduct subscale | 0 | 0 - 1 | 0 | 10 | 99.92 (3.74) | 0.7 | Teacher | 6,365 | 3,588 |
| SDQ conduct subscale | 1 | 0 - 2 | 0 | 10 | 115.82 (1.57) | 0.57 | Mother | 8,069 | 5,541 |
| SDQ conduct subscale | 0 | 0 - 1 | 0 | 10 | 133.99 (3.89) | 0.77 | Teacher | 7,661 | 4,272 |
| SDQ conduct subscale | 1 | 0 - 2 | 0 | 10 | 140.62 (1.64) | 0.59 | Mother | 7,357 | 5,131 |
| SDQ prosocial subscale | 7 | 6 - 9 | 0 | 10 | 48 (1.44) | 1 | Mother | 9,487 | 5,928 |
| SDQ prosocial subscale | 9 | 7 - 10 | 0 | 10 | 81.45 (1.36) | 0.72 | Mother | 8,432 | 5,528 |
| SDQ prosocial subscale | 8 | 7 - 10 | 0 | 10 | 98.43 (3.09) | 1 | Mother | 7,804 | 5,291 |
| SDQ prosocial subscale | 8 | 6 - 10 | 0 | 10 | 99.92 (3.74) | 0.87 | Teacher | 6,364 | 3,586 |
| SDQ prosocial subscale | 9 | 7 - 10 | 0 | 10 | 115.82 (1.57) | 0.68 | Mother | 8,076 | 5,549 |
| SDQ prosocial subscale | 9 | 6 - 10 | 0 | 10 | 133.99 (3.89) | 0.87 | Teacher | 7,666 | 4,273 |
| SDQ prosocial subscale | 9 | 7 - 10 | 0 | 10 | 140.62 (1.64) | 0.69 | Mother | 7,366 | 5,135 |
| SDQ peer problems subscale | 1 | 0 - 2 | 0 | 9 | 48 (1.44) | 0.56 | Mother | 9,499 | 5,931 |
| SDQ peer problems subscale | 1 | 0 - 2 | 0 | 10 | 81.45 (1.36) | 0.59 | Mother | 8,428 | 5,525 |
| SDQ peer problems subscale | 1 | 0 - 2 | 0 | 10 | 98.43 (3.09) | 0.61 | Mother | 7,794 | 5,287 |
| SDQ peer problems subscale | 0 | 0 - 2 | 0 | 10 | 99.92 (3.74) | 0.76 | Teacher | 6,369 | 3,590 |
| SDQ peer problems subscale | 1 | 0 - 2 | 0 | 10 | 115.82 (1.57) | 0.63 | Mother | 8,059 | 5,536 |
| SDQ peer problems subscale | 0 | 0 - 2 | 0 | 10 | 133.99 (3.89) | 0.78 | Teacher | 7,666 | 4,273 |
| SDQ peer problems subscale | 1 | 0 - 2 | 0 | 10 | 140.62 (1.64) | 0.65 | Mother | 7,362 | 5,133 |
| SDQ Internalising subscale | 3 | 1 – 4 | 0 | 16 | 48 (1.44) | 0.7 | Mother | 9,495 | 5,929 |
| SDQ Internalising subscale | 2 | 1 – 4 | 0 | 17 | 81.45 (1.36) | 0.69 | Mother | 8,415 | 5,517 |
| SDQ Internalising subscale | 2 | 1 – 4 | 0 | 19 | 98.43 (3.09) | 0.72 | Mother | 7,789 | 5,285 |
| SDQ Internalising subscale | 1 | 0 – 4 | 0 | 19 | 99.92 (3.74) | 0.82 | Teacher | 6,369 | 3,590 |
| SDQ Internalising subscale | 2 | 1 – 4 | 0 | 19 | 115.82 (1.57) | 0.73 | Mother | 8,032 | 5,516 |
| SDQ Internalising subscale | 1 | 0 – 4 | 0 | 19 | 133.99 (3.89) | 0.84 | Teacher | 7,665 | 4,273 |
| SDQ Internalising subscale | 2 | 1 – 4 | 0 | 20 | 140.62 (1.64) | 0.73 | Mother | 7,333 | 5,116 |
| SDQ Externalising subscale | 6 | 4 – 8 | 0 | 19 | 48 (1.44) | 0.78 | Mother | 9,459 | 5,913 |
| SDQ Externalising subscale | 5 | 2 – 7 | 0 | 19 | 81.45 (1.36) | 0.77 | Mother | 8,392 | 5,507 |
| SDQ Externalising subscale | 4 | 2 – 7 | 0 | 20 | 98.43 (3.09) | 0.80 | Mother | 7,791 | 5,283 |
| SDQ Externalising subscale | 2 | 0 - 5 | 0 | 20 | 99.92 (3.74) | 0.87 | Teacher | 6,360 | 3,585 |
| SDQ Externalising subscale | 4 | 2 – 6 | 0 | 20 | 115.82 (1.57) | 0.77 | Mother | 8,032 | 5,519 |
| SDQ Externalising subscale | 2 | 0 - 5 | 0 | 20 | 133.99 (3.89) | 0.89 | Teacher | 7,661 | 4,272 |
| SDQ Externalising subscale | 3 | 2 – 6 | 0 | 20 | 140.62 (1.64) | 0.78 | Mother | 7,327 | 5,115 |
| Self-Esteem Scholastic | 17 | 15 – 20 | 6 | 24 | 103.83 (3.92) | 0.69 | Child | 6,952 | 5,018 |
| Self-Esteem Global | 20 | 17 – 22 | 6 | 24 | 103.83 (3.92) | 0.65 | Child | 6,941 | 5,011 |

**Legend:** *SDQ Prosocial Score* (this provides a score ranging from 0 to 10, with a higher score indicating better behaviour of the child), *SDQ Hyperactivity Score* (this provides a score ranging from 0 to 10, with a higher score indicating a more hyperactive behaviour of the child), *SDQ Emotional Symptoms Score:* (this provides a score ranging from 0 to 10, with a higher the score indicating more emotional behaviour of the child), *SDQ Conduct Problems Score* (this provides a score ranging from 0 to 10, with a higher score indicating worse behaviour of the child), and *SDQ Peer Problems Score* (this provides a score ranging from 0 to 10, with a higher score indicating that the child behaves in a more pro-social manner with other children). The *SDQ Internalising subscale* is the sum of the Emotional and Peer Problems scales (this provides a score ranging from 0 to 20, with a higher score indicating more internalising problems) and the *SDQ Externalising subscale* is the sum of the Hyperactivity and Conduct Problems scales (this provides a score ranging from 0 to 20, with a higher score indicating more externalising problems).

### Supplementary Table 3. Descriptives of Development and Well-Being Assessment (DAWBA).

| **Variable** | **Median** | | **IQR** | **Min** | | | **Max** | **Mean (SD) age at completion** | | **All Participants with Measure** | | **Genotyped Participants** |
| --- | --- | --- | --- | --- | --- | --- | --- | --- | --- | --- | --- | --- |
| Attention deficit hyperactivity disorder (ADHD) | 0 | 0 – 1 | | | 0 | 5 | | | 91.88 (1.70) | | 8,207 | 5,496 |
| Hyperkinesis | 0 | 0 – 1 | | | 0 | 5 | | | 91.88 (1.70) | | 8,207 | 5,496 |
| Conduct disorder | 1 | 1 – 2 | | | 1 | 5 | | | 91.88 (1.70) | | 8,115 | 5,437 |
| Depressive disorder | 0 | 0 – 1 | | | 0 | 4 | | | 91.88 (1.70) | | 8,089 | 5,427 |
| Generalised anxiety disorder (GAD) | 2 | 1 – 2 | | | 1 | 4 | | | 91.88 (1.70) | | 8,203 | 5,495 |
| Obsessive compulsive disorder (OCD) | 0 | 0 – 0 | | | 0 | 4 | | | 91.88 (1.70) | | 8,234 | 5,512 |
| Oppositional disorder | 2 | 1 – 2 | | | 1 | 5 | | | 91.88 (1.70) | | 8,183 | 5,481 |
| Separation anxiety | 1 | 1 – 1 | | | 1 | 5 | | | 91.88 (1.70) | | 8,215 | 5,502 |
| Separation anxiety (ICD-10) | 0 | 0 – 0 | | | 0 | 4 | | | 91.88 (1.70) | | 8,215 | 5,502 |
| Social phobia | 0 | 0 – 1 | | | 0 | 4 | | | 91.88 (1.70) | | 8,222 | 5,505 |
| Specific phobias | 1 | 0 – 1 | | | 0 | 4 | | | 91.88 (1.70) | | 8,236 | 5,509 |
| Any disorder | 2 | 2 – 2 | | | 1 | 5 | | | 91.88 (1.70) | | 8,252 | 5,519 |
| Any emotional disorder | 2 | 1 – 2 | | | 1 | 5 | | | 91.88 (1.70) | | 8,252 | 5,519 |
| Any anxiety disorder | 2 | 1 – 2 | | | 1 | 5 | | | 91.88 (1.70) | | 8,252 | 5,519 |
| Any behavioural disorder | 2 | 2 – 2 | | | 1 | 5 | | | 91.88 (1.70) | | 8,183 | 5,481 |
| Attention deficit hyperactivity disorder (ADHD) | 0 | 0 – 1 | | | 0 | 5 | | | 128.66 (1.78) | | 7,785 | 5,339 |
| Conduct disorder | 1 | 1 – 2 | | | 1 | 5 | | | 128.66 (1.78) | | 7,687 | 5,282 |
| Depressive disorder | 0 | 0 – 1 | | | 0 | 5 | | | 128.66 (1.78) | | 7,657 | 5,267 |
| Generalised anxiety disorder (GAD) | 2 | 1 – 2 | | | 1 | 4 | | | 128.66 (1.78) | | 7,773 | 5,334 |
| Obsessive compulsive disorder (OCD) | 0 | 0 – 0 | | | 0 | 4 | | | 128.66 (1.78) | | 7,801 | 5,352 |
| Oppositional disorder | 2 | 1 – 2 | | | 1 | 5 | | | 128.66 (1.78) | | 7,759 | 5,326 |
| Separation anxiety | 1 | 1 – 1 | | | 1 | 5 | | | 128.66 (1.78) | | 7,527 | 5,160 |
| Social phobia | 0 | 0 – 1 | | | 0 | 4 | | | 128.66 (1.78) | | 7,797 | 5,347 |
| Specific phobias | 1 | 0 – 1 | | | 0 | 4 | | | 128.66 (1.78) | | 7,788 | 5,338 |
| Any disorder | 2 | 1 – 2 | | | 1 | 5 | | | 128.66 (1.78) | | 7,824 | 5,362 |
| Any emotional disorder | 2 | 1 – 2 | | | 1 | 5 | | | 128.66 (1.78) | | 7,823 | 5,361 |
| Any anxiety disorder | 2 | 1 – 2 | | | 1 | 5 | | | 128.66 (1.78) | | 7,823 | 5,361 |
| Any behavioural disorder | 2 | 1 – 2 | | | 1 | 5 | | | 128.66 (1.78) | | 7,759 | 5,326 |

### Supplementary Table 4. Distribution of socioeconomic, maternal, paternal, familial indicators and child psychosocial indicators in the original ALSPAC cohort in various sub-samples used in the study.

| Variables | | Total ALSPAC-G1 sample (N=14,872) % | Sample with Carey at 6 months and covariates (N= 6,248) % | Sample with Carey at 24 months and covariates (N=6,225) % | Sample with SDQ at 4y and covariates (N= 5,788) % | Sample with SDQ at 6y and covariates (N= 5,435) % | Sample with SDQ at 8y and covariates (N= 5,276) % | Sample with SDQ at 8y* and covariates (N= 3,575) % | Sample with SDQ at 9y and covariates (N=5,488) % | Sample with SDQ at 11y* and covariates (N=4,269) % | Sample with SDQ at 11y and covariates (N= 5,091) % | | Sample with DAWBA at 7y and covariates (N=5,260) % | | Sample with DAWBA at 10y and covariates (N= 4,970) % |
| --- | --- | --- | --- | --- | --- | --- | --- | --- | --- | --- | --- | --- | --- | --- | --- |
| **Maternal education** | | | | | | | | | | | |  | |  | |
|  | A level or higher | 12.89 | 17.71 | 17.05 | 17.15 | 17.26 | 18.61 | 15.71 | 18.44 | 14.57 | 19.25 | | 18.42 | | 19.08 |
|  | O level | 22.38 | 25.87 | 26.23 | 26.36 | 26.81 | 27.44 | 25.68 | 27.05 | 24.88 | 27.20 | | 26.93 | | 27.25 |
|  | <O level | 64.72 | 57.41 | 56.73 | 56.49 | 55,26 | 53.95 | 58.60 | 54.51 | 60.55 | 53.55 | | 54.65 | | 53.67 |
|  | N sample | 12,479 | 6,091 | 6,089 | 5,661 | 5,337 | 5,019 | 3,360 | 5,142 | 3,863 | 4,775 | | 5,129 | | 4,654 |
| **Maternal Social Class** | | | | | | | | | | | | | | | |
|  | High | 37.36 | 42.03 | 42.51 | 42.75 | 43.64 | 44.66 | 40.74 | 44.96 | 39.65 | 44.73 | | 43.84 | | 44.86 |
|  | Low | 62.64 | 57,97 | 57.49 | 57.25 | 56.36 | 55.34 | 59.26 | 55.14 | 60.35 | 55.27 | | 56.16 | | 55.14 |
|  | N sample | 10, 107 | 5,229 | 5,239 | 4,898 | 4,665 | 4,402 | 2,857 | 4,485 | 3,286 | 4,178 | | 4,489 | | 4,070 |
| **Mother smoked in first 3 Months pregnancy** | | | | | | | | | | | | | | | |
|  | No | 74.80 | 80.45 | 80.97 | 80.99 | 81.73 | 82.15 | 80.13 | 82.59 | 78.98 | 82.70 | | 81.92 | | 82.75 |
|  | Yes | 25.20 | 19.55 | 19.03 | 19.01 | 18.27 | 18.85 | 19.87 | 17.41 | 21.02 | 17.30 | | 18.08 | | 17.25 |
|  | N sample | 13,345 | 6,159 | 6,155 | 5,719 | 5,375 | 5,064 | 3,452 | 5,186 | 3,963 | 4,816 | | 5,177 | | 4,708 |

**Legend:** variables with * refer to teacher reported measures.

### Correlations between subscales of SDQ and between mother and teacher reports

Pairwise Pearson correlations between the total score of SDQ across ages varied between 0.47 to 0.73 when reported by the mother and 0.57 when reported by the teacher. Inter-reporter correlations on total SDQ varied between 0.20 to 0.45. Correlation scores for internalising problems ranged from 0.32 to 0.63 when reported by the mother, 0.40 when reported by the teacher and between 0.13 to 0.33 when comparing inter-reporters. Correlation scores for externalising problems ranged from 0.48 to 0.74 when reported by the mother, 0.63 when reported by the teacher and ranged from 0.24 to 0.46 when comparing inter-reporters (**Supplementary Table 5**). Cross-informant correlations were similar to those found by Achenbach et al. ^4^ in a meta-analysis which explored correlations of emotional and behavioural problems of children and adolescents when using multiple reporters.

### Supplementary Table 5. Correlations between SDQ scores across ages and reporters.

| **Variables** | **SDQ_4y** | **SDQ_6y** | **SDQ_8y** | **SDQ_8y*** | **SDQ_9y** | **SDQ_11y*** | **SDQ_11y** |
| --- | --- | --- | --- | --- | --- | --- | --- |
| SDQ_4y | 1 |  |  |  |  |  |  |
| SDQ_6y | 0.58 | 1 |  |  |  |  |  |
| SDQ_8y | 0.57 | 0.73 | 1 |  |  |  |  |
| SDQ_8y* | 0.25 | 0.37 | 0.43 | 1 |  |  |  |
| SDQ_9y | 0.52 | 0.70 | 0.73 | 0.42 | 1 |  |  |
| SDQ_11y* | 0.20 | 0.31 | 0.35 | 0.57 | 0.37 | 1 |  |
| SDQ_11y | 0.47 | 0.64 | 0.67 | 0.42 | 0.72 | 0.43 | 1 |
| **Variables** | **Int_4y** | **Int_6y** | **Int_8y** | **Int_8y*** | **Int_9y** | **Int_11y*** | **Int_11y** |
| Int_4y | 1 |  |  |  |  |  |  |
| Int_6y | 0.46 | 1 |  |  |  |  |  |
| Int_8y | 0.44 | 0.63 | 1 |  |  |  |  |
| Int_8y* | 0.16 | 0.26 | 0.33 | 1 |  |  |  |
| Int_9y | 0.38 | 0.56 | 0.62 | 0.34 | 1 |  |  |
| Int_11y* | 0.13 | 0.22 | 0.27 | 0.40 | 0.31 | 1 |  |
| Int_11y | 0.32 | 0.51 | 0.54 | 0.33 | 0.61 | 0.35 | 1 |
| **Variables** | **Ext_4y** | **Ext_6y** | **Ext_8y** | **Ext_8y*** | **Ext_9y** | **Ext_11y*** | **Ext_11y** |
| Ext_4y | 1 |  |  |  |  |  |  |
| Ext_6y | 0.59 | 1 |  |  |  |  |  |
| Ext_8y | 0.58 | 0.73 | 1 |  |  |  |  |
| Ext_8y* | 0.28 | 0.42 | 0.46 | 1 |  |  |  |
| Ext_9y | 0.54 | 0.71 | 0.74 | 0.45 | 1 |  |  |
| Ext_11y* | 0.24 | 0.34 | 0.38 | 0.63 | 0.39 | 1 |  |
| Ext_11y | 0.48 | 0.65 | 0.69 | 0.45 | 0.73 | 0.45 | 1 |

**Legend:** SDQ_4y to SDQ_11y correspond to the total score on the SDQ; Int_4y to Int_11y correspond to the internalising scale of the SDQ (emotional subscale + peer problems subscale); Ext_4y to Ext_11y correspond to the externalising scale of the SDQ (hyperactivity subscale + conduct problems subscale).

Asterisk* indicates when the SDQ was reported by the teacher.

### Supplementary Table 6. Separate linear regressions of child PRS toward neuroticism on various emotional and behavioural outcomes. Estimated with robust standard errors adjusted for sex, age and the first 5 principal components of genetic ancestry.

| **Outcome variable** | **Age** | **Report** | **N** | **Beta** | **95% Confidence Intervals** | **P-value** | **R^2^** |
| --- | --- | --- | --- | --- | --- | --- | --- |
| Emotional Stability (Big 5) | 13 years | Child-self report | 4224 | -0.75 | (-0.96, -0.55) | 7.460 x 10^-13^ | 0.07 |
| Prorated score of Carey – Activity subscale | 6 months | Mother | 6271 | -0.16 | (-0.33, 0.00) | 0.057 | 0.01 |
| Prorated score of Carey – Rhythm subscale | 6 months | Mother | 6269 | 0.08 | (-0.10, 0.26) | 0.393 | 0.001 |
| Prorated score of Carey – Approach subscale | 6 months | Mother | 6266 | 0.18 | (0.01, 0.35) | 0.034 | 0.01 |
| Prorated score of Carey – Adaptability subscale | 6 months | Mother | 6271 | 0.11 | (-0.04, 0.27) | 0.139 | 0.003 |
| Prorated score of Carey – Intensity subscale | 6 months | Mother | 6269 | 0.01 | (-0.14, 0.16) | 0.896 | 0.001 |
| Prorated score of Carey – Mood subscale | 6 months | Mother | 6267 | 0.03 | (-0.12, 0.19) | 0.665 | 0.001 |
| Prorated score of Carey – Persistence subscale | 6 months | Mother | 6269 | -0.03 | (-0.16, 0.11) | 0.716 | 0.002 |
| Prorated score of Carey – Distract subscale | 6 months | Mother | 6271 | 0.07 | (-0.07, 0.22) | 0.323 | 0.001 |
| Prorated score of Carey – Threshold | 6 months | Mother | 6264 | 0.21 | (0.05, 0.37) | 0.010 | 0.004 |
| Prorated score of Carey – Activity subscale | 24 months | Mother | 6260 | 0.08 | (-0.04, 0.20) | 0.174 | 0.01 |
| Prorated score of Carey – Rhythm subscale | 24 months | Mother | 6257 | 0.25 | (0.09, 0.40) | 0.002 | 0.003 |
| Prorated score of Carey – Approach subscale | 24 months | Mother | 6253 | 0.17 | (-0.03, 0.36) | 0.092 | 0.01 |
| Prorated score of Carey – Adaptability subscale | 24 months | Mother | 6233 | 0.15 | (0.04, 0.27) | 0.006 | 0.01 |
| Prorated score of Carey – Intensity subscale | 24 months | Mother | 6255 | 0.14 | (0.02, 0.26) | 0.027 | 0.002 |
| Prorated score of Carey – Mood subscale | 24 months | Mother | 6259 | 0.21 | (0.06, 0.36) | 0.005 | 0.003 |
| Prorated score of Carey – Persistence subscale | 24 months | Mother | 6252 | 0.20 | (0.07, 0.32) | 0.002 | 0.01 |
| Prorated score of Carey – Distract subscale | 24 months | Mother | 6255 | 0.07 | (-0.05, 0.20) | 0.260 | 0.001 |
| Prorated score of Carey – Threshold | 24 months | Mother | 6260 | -0.10 | (-0.21, 0.02) | 0.113 | 0.02 |
| Prosocial prorated subscale of SDQ | 4 years | Mother | 5803 | -0.04 | (-0.10, 0.01) | 0.117 | 0.02 |
| Hyperactive prorated subscale of SDQ | 4 years | Mother | 5808 | 0.06 | (-0.01, 0.12) | 0.074 | 0.02 |
| Emotional prorated subscale of SDQ | 4 years | Mother | 5807 | 0.05 | (0.01, 0.09) | 0.014 | 0.002 |
| Conduct prorated subscale of SDQ | 4 years | Mother | 5803 | 0.08 | (0.04, 0.11) | 7.46 x 10^-6^ | 0.01 |
| Peer problems prorated subscale of SDQ | 4 years | Mother | 5806 | 0.09 | (0.05, 0.13) | 1.03 x 10^-5^ | 0.01 |
| Total prorated score of SDQ | 4 years | Mother | 5788 | 0.27 | (0.15, 0.40) | 2.30 x 10^-5^ | 0.02 |
| Prosocial prorated subscale of SDQ | 6 years | Parent | 5524 | -0.01 | (-0.06, 0.04) | 0.59 | 0.04 |
| Hyperactive prorated subscale of SDQ | 6 years | Parent | 5518 | 0.08 | (0.02, 0.15) | 0.011 | 0.03 |
| Emotional prorated subscale of SDQ | 6 years | Parent | 5517 | 0.12 | (0.08, 0.17) | 1.73 x 10^-7^ | 0.01 |
| Conduct prorated subscale of SDQ | 6 years | Parent | 5525 | 0.1 | (0.06, 0.14) | 9.58 x 10^-7^ | 0.01 |
| Peer problems prorated subscale of SDQ | 6 years | Parent | 5521 | 0.08 | (0.04, 0.12) | 8.85 x 10^-5^ | 0.01 |
| Total prorated score of SDQ | 6 years | Parent | 5510 | 0.37 | (0.24, 0.50) | 2.43 x 10^-8^ | 0.02 |
| Prosocial prorated subscale of SDQ | 8 years | Mother | 5287 | -0.08 | (-0.14, -0.03) | 0.003 | 0.03 |
| Hyperactive prorated subscale of SDQ | 8 years | Mother | 5282 | 0.12 | (0.05, 0.19) | 0.001 | 0.04 |
| Emotional prorated subscale of SDQ | 8 years | Mother | 5282 | 0.17 | (0.12, 0.23) | 1.38 x 10^-9^ | 0.01 |
| Conduct prorated subscale of SDQ | 8 years | Mother | 5283 | 0.12 | (0.08, 0.17) | 1.73 x 10^-7^ | 0.01 |
| Peer problems prorated subscale of SDQ | 8 years | Mother | 5283 | 0.10 | (0.06, 0.14) | 1.33 x 10^-5^ | 0.01 |
| Total prorated score of SDQ | 8 years | Mother | 5277 | 0.52 | (0.38, 0.67) | 2.08 x 10^-12^ | 0.03 |
| Prosocial prorated subscale of SDQ | 8 years | Teacher | 3579 | -0.13 | (-0.21, -0.05) | 0.001 | 0.10 |
| Hyperactive prorated subscale of SDQ | 8 years | Teacher | 3583 | 0.17 | (0.09, 0.26) | 8.85 x 10^-5^ | 0.10 |
| Emotional prorated subscale of SDQ | 8 years | Teacher | 3583 | 0.14 | (0.07, 0.21) | 8.85 x 10^-5^ | 0.01 |
| Conduct prorated subscale of SDQ | 8 years | Teacher | 3581 | 0.1 | (0.05, 0.14) | 1.33 x 10^-5^ | 0.05 |
| Peer problems prorated subscale of SDQ | 8 years | Teacher | 3583 | 0.1 | (0.04, 0.16) | 0.001 | 0.02 |
| Total prorated score of SDQ | 8 years | Teacher | 3582 | 0.52 | (0.33, 0.71) | 8.13 x 10^-8^ | 0.07 |
| Prosocial prorated subscale of SDQ | 9 years | Parent | 5545 | -0.04 | (-0.09, 0.00) | 0.073 | 0.04 |
| Hyperactive prorated subscale of SDQ | 9 years | Parent | 5538 | 0.1 | (0.04, 0.16) | 0.001 | 0.04 |
| Emotional prorated subscale of SDQ | 9 years | Parent | 5524 | 0.14 | (0.09, 0.19) | 4.07 x 10^-8^ | 0.01 |
| Conduct prorated subscale of SDQ | 9 years | Parent | 5537 | 0.1 | (0.06, 0.14) | 9.58 x 10^-7^ | 0.01 |
| Peer problems prorated subscale of SDQ | 9 years | Parent | 5532 | 0.1 | (0.06, 0.15) | 1.33 x 10^-5^ | 0.01 |
| Total prorated score of SDQ | 9 years | Parent | 5516 | 0.45 | (0.31, 0.59) | 2.98 x 10^-10^ | 0.02 |
| Prosocial prorated subscale of SDQ | 11 years | Teacher | 4270 | -0.06 | (-0.13, 0.02) | 0.128 | 0.10 |
| Hyperactive prorated subscale of SDQ | 11 years | Teacher | 4270 | 0.09 | (0.02, 0.17) | 0.019 | 0.12 |
| Emotional prorated subscale of SDQ | 11 years | Teacher | 4270 | 0.09 | (0.03, 0.15) | 0.002 | 0.01 |
| Conduct prorated subscale of SDQ | 11 years | Teacher | 4269 | 0.08 | (0.03, 0.13) | 0.001 | 0.06 |
| Peer problems prorated subscale of SDQ | 11 years | Teacher | 4270 | 0.07 | (0.02, 0.13) | 0.011 | 0.01 |
| Total prorated score of SDQ | 11 years | Teacher | 4270 | 0.34 | (0.17, 0.51) | 8.85 x 10^-5^ | 0.07 |
| Prosocial prorated subscale of SDQ | 11 years | Parent | 5132 | -0.02 | (-0.06, 0.03) | 0.507 | 0.04 |
| Hyperactive prorated subscale of SDQ | 11 years | Parent | 5118 | 0.11 | (0.05, 0.17) | 0.001 | 0.04 |
| Emotional prorated subscale of SDQ | 11 years | Parent | 5118 | 0.18 | (0.13, 0.23) | 1.71 x 10^-12^ | 0.02 |
| Conduct prorated subscale of SDQ | 11 years | Parent | 5128 | 0.09 | (0.05, 0.13) | 1.03 x 10^-5^ | 0.01 |
| Peer problems prorated subscale of SDQ | 11 years | Parent | 5130 | 0.1 | (0.05, 0.14) | 1.33 x 10^-5^ | 0.01 |
| Total prorated score of SDQ | 11 years | Parent | 5128 | 0.47 | (0.33, 0.61) | 4.70 x 10^-11^ | 0.02 |
| Locus of control | 8 years | Child | 4599 | 0.13 | (0.07, 0.19) | 2.17 x 10^-5^ | 0.01 |
| Self-esteem global score | 8 years | Child | 5009 | -0.13 | (-0.24, -0.03) | 0.01 | 0.003 |
| Self-Esteem scholastic | 8 years | Child | 5016 | -0.21 | (-0.31, -0.10) | 8.85 x 10^-5^ | 0.01 |
| IQ measured with WISC | 8 years | Child | 5295 | -1.15 | (-1.61, -0.68) | 1.25 x 10^-6^ | 0.02 |

### Supplementary Table 7. Separate ordinal regressions of a child PRS for neuroticism on various emotional and behavioural outcomes.

| **Outcome variable**  **(DAWBA)** | **Age** | **Report** | **Number** | **OR** | **95% Confidence Intervals** | **P-value** | **R^2^** |
| --- | --- | --- | --- | --- | --- | --- | --- |
| Attention deficit hyperactivity disorder (ADHD) | 7 years | Parent and teacher | 5448 | 1.09 | (1.04, 1.16) | 0.004 | 0.02 |
| Conduct disorder | 7 years | Parent and teacher | 5389 | 1.06 | (1.00, 1.13) | 0.036 | 0.003 |
| Depressive disorder | 7 years | Parent and teacher | 5379 | 1.05 | (0.99, 1.12) | 0.074 | 0.001 |
| Generalised anxiety disorder (GAD) | 7 years | Parent and teacher | 5446 | 1.13 | (1.06, 1.20) | 8.85 x 10^-5^ | 0.004 |
| Obsessive compulsive disorder (OCD) | 7 years | Parent and teacher | 5463 | 1.08 | (1.00, 1.16) | 0.042 | 0.002 |
| Oppositional disorder | 7 years | Parent and teacher | 5432 | 1.09 | (1.03, 1.15) | 0.002 | 0.01 |
| Separation anxiety | 7 years | Parent and teacher | 5453 | 1.17 | (1.07, 1.27) | 0.0002 | 0.001 |
| Separation anxiety (ICD-10) | 7 years | Parent and teacher | 5453 | 1.23 | (1.11, 1.38) | 0.0002 | 0.01 |
| Social phobia | 7 years | Parent and teacher | 5456 | 1.06 | (1.00, 1.14) | 0.054 | 0.003 |
| Specific phobias | 7 years | Parent and teacher | 5460 | 1.09 | (1.04, 1.16) | 0.001 | 0.004 |
| Any disorder | 7 years | Parent and teacher | 5470 | 1.14 | (1.06, 1.21) | 8.85 x 10^-5^ | 0.01 |
| Any emotional disorder | 7 years | Parent and teacher | 5470 | 1.14 | (1.07, 1.20) | 3.61 x 10^-6^ | 0.004 |
| Any anxiety disorder | 7 years | Parent and teacher | 5470 | 1.13 | (1.07, 1.20) | 1.90 x 10^-5^ | 0.004 |
| Any behavioural disorder | 7 years | Parent and teacher | 5432 | 1.09 | (1.04, 1.16) | 0.001 | 0.01 |
| Attention deficit hyperactivity disorder (ADHD) | 10 years | Parent | 5336 | 1.11 | (1.04, 1.17) | 0.002 | 0.01 |
| Conduct disorder | 10 years | Parent | 5279 | 1.03 | (0.97, 1.09) | 0.343 | 0.003 |
| Depressive disorder | 10 years | Parent | 5264 | 1.11 | (1.04, 1.17) | 0.001 | 0.01 |
| Generalised anxiety disorder (GAD) | 10 years | Parent | 5331 | 1.14 | (1.08, 1.21) | 3.61 x 10^-6^ | 0.004 |
| Obsessive compulsive disorder (OCD) | 10 years | Parent | 5349 | 1.14 | (1.04, 1.24) | 0.002 | 0.003 |
| Oppositional disorder | 10 years | Parent | 5323 | 1.09 | (1.04, 1.16) | 0.001 | 0.01 |
| Separation anxiety | 10 years | Parent | 5157 | 1.21 | (1.11, 1.31) | 1.1 x 10^-5^ | 0.01 |
| Social phobia | 10 years | Parent | 5344 | 1.12 | (1.04, 1.19) | 0.001 | 0.002 |
| Specific phobias | 10 years | Parent | 5335 | 1.13 | (1.06, 1.20) | 8.85 x 10^-5^ | 0.01 |
| Any disorder | 10 years | Parent | 5359 | 1.17 | (1.09, 1.25) | 1.40 x 10^-6^ | 0.01 |
| Any emotional disorder | 10 years | Parent | 5358 | 1.16 | (1.09, 1.23) | 9.58 x 10^-7^ | 0.01 |
| Any anxiety disorder | 10 years | Parent | 5358 | 1.19 | (1.12, 1.26) | 2.80 x 10^-8^ | 0.004 |
| Any behavioural disorder | 10 years | Parent | 5323 | 1.09 | (1.03, 1.16) | 0.003 | 0.01 |

**Legend:** Ordinal regressions estimated with robust SEs adjusted for sex, age and the first 5 principal components of genetic ancestry.

### Sensitivity analyses: exploring differential misclassification depending on maternal PRS

Differential misclassification is a systematic bias in the measurement of the outcome depending on a third factor. Here, we hypothesised that mothers with a higher PRS for neuroticism may have been more sensitive to negative emotions in the child compared to women with a lower liability to neuroticism, leading to a higher score on scales measuring emotional and behavioural problems of the child. We employed different methods to explore the presence of differential misclassification of the child outcome (i.e., SDQ) dependent on a maternal PRS for neuroticism. We first used maternal genotype (N=7,826) as the exposure when assessing child’s behaviour as reported by the child’s teacher as an outcome while adjusting for child genotype. No evidence of association between the maternal PRS for neuroticism and the teacher rating of child behaviours, once adjusting for the child PRS for neuroticism, would suggest that the teacher rating is independent of the maternal PRS. Second, because of the availability of both maternal and teacher scoring of the SDQ at two different timepoints at approximately the same age (around 8 and 11 years of age), we explored the potential effect of differential misclassification dependent on the maternal PRS for neuroticism by generating a variable which represented the difference in score between the teacher and mother reported scores of child behaviours. This variable was obtained by subtracting the maternally reported score of child’s problems (measured with the SDQ at 8 and 11 years of age) from the teacher reported score of child’s problems (measured with the SDQ at 8 and 11 years of age). If this score had a mean value of 0, it suggested that no difference was present in the rating performed by the mother as compared to the teacher.

However, simply observing a difference in score does not imply that differential misclassification occurred. In fact, the difference could be representative of a real difference in the child behaviour (e.g., the child is clingier and less hyperactive at home than he or she is at school or vice-versa), resulting in a differential assessment by the parent and the teacher. However, if the difference in score was uniquely induced by a true difference in behaviour, we would expect it to be independent of the maternal PRS once adjusting for the child’s own PRS. Thus, we employed linear regression models adjusting for the child PRS to explore the association between the maternal PRS and the difference in score.

### Supplementary Table 8. Associations between maternal neuroticism PRS and child’s emotional and behavioural difficulties as rated by the teacher adjusted for the child’s neuroticism PRS.

| **Outcome** | **Number** | **Beta** | **95% CIs** | **P-values** |
| --- | --- | --- | --- | --- |
| SDQ_prosoc_8y | 2389 | -0.05 | (-0.15,0.06) | 0.375 |
| SDQ_hyperac_8y | 2392 | 0.06 | (-0.06,0.17) | 0.312 |
| SDQ_emot_8y | 2392 | 0.03 | (-0.06,0.12) | 0.533 |
| SDQ_cond_8y | 2391 | -0.00 | (-0.06,0.06) | 0.963 |
| SDQ_peer_8y | 2392 | 0.02 | (-0.07,0.10) | 0.715 |
| SDQ_total_8y | 2392 | 0.10 | (-0.16,0.35) | 0.456 |
| SDQ_prosoc_11y | 2841 | -0.04 | (-0.14,0.05) | 0.358 |
| SDQ_hyperac_11y | 2841 | 0.02 | (-0.08,0.12) | 0.693 |
| SDQ_emot_11y | 2841 | 0.04 | (-0.04,0.11) | 0.314 |
| SDQ_cond_11y | 2841 | -0.01 | (-0.07,0.05) | 0.800 |
| SDQ_peer_11y | 2841 | 0.03 | (-0.05,0.11) | 0.448 |
| SDQ_total_11y | 2841 | 0.08 | (-0.15,0.31) | 0.487 |

**Legend:** All outcomes are reported by the teacher. Separate linear regression with robust SEs were employed. Analyses were adjusted for child’s sex, age, PRS, and the first five principal components of genetic ancestry.

### Supplementary Table 9. Descriptives of “difference in score” variables.

| **Variable** | **Number** | **Mean** | **SD** | **Min** | **Max** |
| --- | --- | --- | --- | --- | --- |
| Prosocial difference score (teacher 8y – mother 8y) | 3,685 | -0.20 | 2.64 | -9 | 8 |
| Hyperactivity difference score (teacher 8y – mother 8y) | 3,682 | -0.93 | 2.60 | -10 | 9 |
| Emotional difference score (teacher 8y – mother 8y) | 3,683 | -0.38 | 2.22 | -9 | 9 |
| Conduct difference score (teacher 8y – mother 8y) | 3,683 | -0.84 | 1.56 | -8 | 8 |
| Peer problems difference score (teacher 8y – mother 8y) | 3,684 | -0.19 | 1.83 | -9 | 10 |
| Total difference score (teacher 8y – mother 8y) | 3,678 | -2.35 | 5.50 | -28 | 21 |
| Prosocial difference score (teacher 11y – mother 11y) | 4,144 | -0.25 | 2.54 | -9 | 10 |
| Hyperactivity difference score (teacher 11y – mother 11y) | 4,123 | -0.81 | 2.47 | -10 | 9 |
| Emotional difference score (teacher 11y – mother 11y) | 4,132 | -0.28 | 2.15 | -9 | 9 |
| Conduct difference score (teacher 11y – mother 11y) | 4,143 | -0.56 | 1.62 | -9 | 9 |
| Peer problems difference score (teacher 11y – mother 11y) | 4,147 | -0.00 | 1.84 | -9 | 10 |
| Total difference score (teacher 11y – mother 11y) | 4,138 | -1.64 | 5.46 | -29 | 25 |

**Legend:** Child’s scores on the SDQ subscales and total score as rated by the mother were subtracted from the child’s score on the SDQ as rated by the teacher at similar ages. Negative means indicate higher SDQ scores as rated by the mother.

### Supplementary Table 10. Associations between maternal neuroticism PRS and “difference in score” outcomes (teacher - mother reported scores) unadjusted and adjusted for child neuroticism PRS.

|  |  | **Unadjusted complete case** | | **Adjusted for child PRS** | |
| --- | --- | --- | --- | --- | --- |
| **Outcome (difference in scores)** | **Number** | **Beta** | **95% CIs** | **Beta** | **95% CIs** |
| Prosocial (teacher 8y – mother 8y) | 1805 | -0.05 | (-0.17,0.07) | -0.10 | (-0.24,0.04) |
| Hyperactivity (teacher 8y – mother 8y) | 1805 | 0.06 | (-0.06,0.18) | -0.03 | (-0.17,0.10) |
| Emotional (teacher 8y – mother 8y) | 1807 | 0.06 | (-0.04,0.16) | 0.10 | (-0.01,0.22) |
| Conduct (teacher 8y – mother 8y) | 1806 | 0.03 | (-0.04,0.10) | 0.04 | (-0.04,0.13) |
| Peer problems (teacher 8y – mother 8y) | 1807 | 0.05 | (-0.03,0.14) | 0.08 | (-0.02,0.18) |
| Total score (teacher 8y – mother 8y) | 1804 | 0.19 | (-0.06,0.44) | 0.29 | (0.00,0.58) |
| Prosocial (teacher 11y – mother 11y) | 2104 | 0.02 | (-0.08,0.13) | -0.02 | (-0.14,0.10) |
| Hyperactivity (teacher 11y – mother 11y) | 2096 | -0.09 | (-0.19,0.01) | -0.11 | (-0.22,0.01) |
| Emotional (teacher 11y – mother 11y) | 2098 | -0.08 | (-0.16,0.01) | -0.04 | (-0.14,0.06) |
| Conduct (teacher 11y – mother 11y) | 2105 | -0.06 | (-0.12,0.01) | -0.06 | (-0.14,0.02) |
| Peer problems (teacher 11y – mother 11y) | 2104 | -0.03 | (-0.10,0.05) | 0.01 | (-0.08,0.10) |
| Total score (teacher 11y – mother 11y) | 2102 | -0.25 | (-0.48, -0.03) | -0.20 | (-0.46,0.06) |

**Legend:** Separate linear regression with robust SEs were employed. Analyses were adjusted for child’s sex, age, PRS, and the first five principal components of genetic ancestry. SDQ subscales and total score at 8 and 11 years of age when reported by both mothers and teachers.

### Sensitivity analyses: exploring the causal nature of primary analyses

To address one of the secondary aims of this study (i.e., estimating the causal relationship between the child neuroticism PRS and later emotional and behavioural difficulties) we adjusted our analyses for the maternal PRS for neuroticism in the separate multivariable linear regressions in order to account for a possible confounding effect of this variable. Paternal PRS was not employed in models for two main reasons: first, we lacked statistical power as only 1,711 fathers had genetic information available for this analysis. Second, as suggested in prior studies ^5^, maternal and paternal genetics may have differential effects on the child’s phenotype. Here, we hypothesised that the greatest variance in child’s genotype and phenotype would have been contributed by maternal genotype via a greater “genetic nurturing” effect.

### Supplementary Table 11. Associations between neuroticism PRS and all psychological outcomes, unadjusted and adjusted for maternal neuroticism PRS.

|  | | | | **Complete Unadjusted for maternal PRS** | | | **Complete adjusted for maternal PRS** | | |
| --- | --- | --- | --- | --- | --- | --- | --- | --- | --- |
| **Outcome variable (Linear regression models)** | **Age** | **Report** | **Number** | **Beta** | **95% CIs** | **P-values** | **Beta** | **95% CIs** | **P-values** |
| Prorated score of Carey – Activity subscale | 6 months | Mother | 4271 | -0.12 | (-0.32,0.09) | 0.27 | -0.12 | (-0.35,0.11) | 0.30 |
| Prorated score of Carey – Rhythm subscale | 6 months | Mother | 4271 | 0.04 | (-0.19,0.26) | 0.76 | 0.04 | (-0.21,0.29) | 0.78 |
| Prorated score of Carey – Approach subscale | 6 months | Mother | 4268 | 0.25 | (0.05,0.45) | 0.02 | 0.27 | (0.05,0.50) | 0.02 |
| Prorated score of Carey – Adaptability subscale | 6 months | Mother | 4272 | 0.18 | (-0.00,0.36) | 0.06 | 0.27 | (0.07,0.47) | 0.01 |
| Prorated score of Carey – Intensity subscale | 6 months | Mother | 4270 | 0.09 | (-0.10,0.27) | 0.35 | 0.07 | (-0.14,0.28) | 0.51 |
| Prorated score of Carey – Mood subscale | 6 months | Mother | 4269 | 0.08 | (-0.10,0.27) | 0.380 | 0.18 | (-0.04,0.39) | 0.105 |
| Prorated score of Carey – Persistence subscale | 6 months | Mother | 4270 | -0.05 | (-0.21,0.11) | 0.55 | 0.07 | (-0.12,0.25) | 0.47 |
| Prorated score of Carey – Distract subscale | 6 months | Mother | 4271 | 0.15 | (-0.03,0.33) | 0.10 | 0.20 | (0.00,0.40) | 0.05 |
| Prorated score of Carey – Threshold | 6 months | Mother | 4269 | 0.22 | (0.03,0.41) | 0.02 | 0.15 | (-0.06,0.37) | 0.16 |
| Prorated score of Carey – Activity subscale | 24 months | Mother | 4241 | 0.15 | (0.01,0.29) | 0.03 | 0.16 | (0.00,0.32) | 0.05 |
| Prorated score of Carey – Rhythm subscale | 24 months | Mother | 4238 | 0.31 | (0.12,0.49) | 2.276 x 10^-8^ | 0.26 | (0.04,0.47) | 0.02 |
| Prorated score of Carey – Approach subscale | 24 months | Mother | 4235 | 0.10 | (-0.14,0.35) | 0.40 | 0.02 | (-0.26,0.30) | 0.89 |
| Prorated score of Carey – Adaptability subscale | 24 months | Mother | 4229 | 0.19 | (0.06,0.33) | 0.01 | 0.17 | (0.02,0.32) | 0.03 |
| Prorated score of Carey – Intensity subscale | 24 months | Mother | 4237 | 0.18 | (0.03,0.33) | 0.02 | 0.20 | (0.02,0.37) | 0.03 |
| Prorated score of Carey – Mood subscale | 24 months | Mother | 4241 | 0.29 | (0.11,0.47) | 2.276 x 10^-8^ | 0.30 | (0.09,0.50) | 0.004 |
| Prorated score of Carey – Persistence subscale | 24 months | Mother | 4235 | 0.21 | (0.05,0.36) | 0.01 | 0.22 | (0.04,0.40 | 0.02 |
| Prorated score of Carey – Distract subscale | 24 months | Mother | 4238 | 0.04 | (-0.11,0.19) | 0.60 | 0.07 | (-0.10,0.24) | 0.44 |
| Prorated score of Carey – Threshold | 24 months | Mother | 4241 | -0.10 | (-0.25,0.04) | 0.17 | -0.15 | (-0.31,0.02) | 0.08 |
| Prosocial prorated subscale of SDQ | 4 years | Mother | 3990 | -0.06 | (-0.12,0.01) | 0.08 | -0.04 | (-0.12,0.03) | 0.24 |
| Hyperactive prorated subscale of SDQ | 4 years | Mother | 3996 | 0.09 | (0.01,0.16) | 0.02 | 0.06 | (-0.03,0.14) | 0.18 |
| Emotional prorated subscale of SDQ | 4 years | Mother | 3996 | 0.08 | (0.03,0.13) | 0.003 | 0.07 | (0.01,0.12) | 0.02 |
| Conduct prorated subscale of SDQ | 4 years | Mother | 3990 | 0.10 | (0.05,0.14) | 2.276 x 10^-8^ | 0.07 | (0.02,0.13) | 0.01 |
| Peer problems prorated subscale of SDQ | 4 years | Mother | 3994 | 0.09 | (0.04,0.14) | 0.0004 | 0.05 | (-0.00,0.11) | 0.06 |
| Total prorated score of SDQ | 4 years | Mother | 3980 | 0.35 | (0.20,0.50) | 1.327 x 10^-5^ | 0.25 | (0.08,0.42) | 0.003 |
| Prosocial prorated subscale of SDQ | 6 years | Mother | 3805 | -0.02 | (-0.08,0.03) | 0.41 | -0.05 | (-0.11,0.02) | 0.14 |
| Hyperactive prorated subscale of SDQ | 6 years | Mother | 3802 | 0.07 | (-0.01,0.15) | 0.07 | 0.04 | (-0.05,0.13) | 0.35 |
| Emotional prorated subscale of SDQ | 6 years | Mother | 3801 | 0.12 | (0.07,0.17) | 2.55 x 10^-6^ | 0.10 | (0.04,0.17) | 0.001 |
| Conduct prorated subscale of SDQ | 6 years | Mother | 3804 | 0.09 | (0.04,0.14) | 0.0004 | 0.09 | (0.03,0.14) | 0.002 |
| Peer problems prorated subscale of SDQ | 6 years | Mother | 3801 | 0.08 | (0.03,0.12) | 0.001 | 0.07 | (0.02,0.13) | 0.006 |
| Total prorated score of SDQ | 6 years | Mother | 3796 | 0.34 | (0.19,0.50) | 1.71 x 10^-5^ | 0.29 | (0.11,0.47) | 0.002 |
| Prosocial prorated subscale of SDQ | 8 years | Mother | 3668 | -0.08 | (-0.14, -0.01) | 0.02 | -0.07 | (-0.14, -0.00) | 0.05 |
| Hyperactive prorated subscale of SDQ | 8 years | Mother | 3665 | 0.11 | (0.03,0.20) | 0.01 | 0.08 | (-0.02,0.17) | 0.11 |
| Emotional prorated subscale of SDQ | 8 years | Mother | 3666 | 0.19 | (0.13,0.25) | 5.41 x 10^-10^ | 0.17 | (0.10,0.25) | 8.89 x 10^-6^ |
| Conduct prorated subscale of SDQ | 8 years | Mother | 3666 | 0.12 | (0.07,0.17) | 2.55 x 10^-6^ | 0.11 | (0.05,0.17) | 0.0003 |
| Peer problems prorated subscale of SDQ | 8 years | Mother | 3666 | 0.10 | (0.05,0.15) | 8.86 x 10^-5^ | 0.09 | (0.03,0.15) | 0.003 |
| Total prorated score of SDQ | 8 years | Mother | 3663 | 0.53 | (0.36,0.71) | 2.92 x 10^-9^ | 0.45 | (0.25,0.66) | 1.69 x 10^-5^ |
| Prosocial prorated subscale of SDQ | 8 years | Teacher | 2389 | -0.12 | (-0.22, -0.02) | 0.02 | -0.10 | (-0.21,0.01) | 0.09 |
| Hyperactive prorated subscale of SDQ | 8 years | Teacher | 2392 | 0.14 | (0.03,0.24) | 0.01 | 0.11 | (-0.01,0.23) | 0.08 |
| Emotional prorated subscale of SDQ | 8 years | Teacher | 2392 | 0.13 | (0.05,0.21) | 0.003 | 0.12 | (0.02,0.21) | 0.02 |
| Conduct prorated subscale of SDQ | 8 years | Teacher | 2391 | 0.09 | (0.04,0.14) | 0.001 | 0.09 | (0.03,0.15) | 0.01 |
| Peer problems prorated subscale of SDQ | 8 years | Teacher | 0.11;(0.04,0.18) | 0.11 | (0.04,0.18) | 0.002 | 0.10 | (0.02,0.19) | 0.02 |
| Total prorated score of SDQ | 8 years | Teacher | 2392 | 0.47 | (0.25,0.70) | 4.24 x 10^-5^ | 0.42 | (0.16,0.69) | 0.002 |
| Prosocial prorated subscale of SDQ | 9 years | Mother | 3811 | -0.05 | (-0.10,0.01) | 0.09 | -0.05 | (-0.11,0.01) | 0.13 |
| Hyperactive prorated subscale of SDQ | 9 years | Mother | 3805 | 0.09 | (0.02,0.16) | 0.02 | 0.05 | (-0.04,0.13) | 0.26 |
| Emotional prorated subscale of SDQ | 9 years | Mother | 3802 | 0.13 | (0.07,0.19) | 2.17 x 10^-5^ | 0.09 | (0.03,0.16) | 0.004 |
| Conduct prorated subscale of SDQ | 9 years | Mother | 3806 | 0.09 | (0.04,0.14) | 0.0004 | 0.07 | (0.02,0.12) | 0.01 |
| Peer problems prorated subscale of SDQ | 9 years | Mother | 3802 | 0.12 | (0.07,0.17) | 2.55 x 10^-6^ | 0.09 | (0.03,0.15) | 0.002 |
| Total prorated score of SDQ | 9 years | Mother | 3795 | 0.43 | (0.27,0.59) | 1.38 x 10^-7^ | 0.30 | (0.12,0.49) | 0.002 |
| Prosocial prorated subscale of SDQ | 11 years | Teacher | 2841 | -0.05 | (-0.14,0.03) | 0.24 | -0.03 | (-0.13,0.07) | 0.54 |
| Hyperactive prorated subscale of SDQ | 11 years | Teacher | 2841 | 0.09 | (-0.00,0.19) | 0.06 | 0.08 | (-0.03,0.19) | 0.14 |
| Emotional prorated subscale of SDQ | 11 years | Teacher | 2841 | 0.07 | (0.00,0.14) | 0.04 | 0.05 | (-0.02,0.13) | 0.17 |
| Conduct prorated subscale of SDQ | 11 years | Teacher | 2841 | 0.06 | (0.01,0.12) | 0.03 | 0.07 | (0.00,0.13) | 0.04 |
| Peer problems prorated subscale of SDQ | 11 years | Teacher | 2841 | 0.05 | (-0.02,0.12) | 0.19 | 0.03 | (-0.05,0.11) | 0.44 |
| Total prorated score of SDQ | 11 years | Teacher | 2841 | 0.27 | (0.06,0.49) | 0.01 | 0.23 | (-0.01,0.48) | 0.06 |
| Prosocial prorated subscale of SDQ | 11 years | Mother | 3621 | -0.04 | (-0.09,0.02) | 0.20 | -0.03 | (-0.09,0.04) | 0.42 |
| Hyperactive prorated subscale of SDQ | 11 years | Mother | 3613 | 0.88 | (0.74,1.02) | 7.07 x 10^-35^ | 0.04 | (-0.05,0.12) | 0.37 |
| Emotional prorated subscale of SDQ | 11 years | Mother | 3614 | 0.18 | (0.12,0.23) | 1.41 x 10^-10^ | 0.15 | (0.08,0.22) | 2.67 x 10^-5^ |
| Conduct prorated subscale of SDQ | 11 years | Mother | 3622 | 0.06 | (0.01,0.11) | 0.02 | 0.03 | (-0.02,0.09) | 0.24 |
| Peer problems prorated subscale of SDQ | 11 years | Mother | 3622 | 0.10 | (0.04,0.15) | 0.0004 | 0.08 | (0.02,0.14) | 0.009 |
| Total prorated score of SDQ | 11 years | Mother | 3620 | 0.40 | (0.23,0.57) | 3.99 x 10^-6^ | 0.29 | (0.10,0.49) | 0.003 |
| Locus of control | 8 years | Child | 3796 | 0.16 | (0.05,0.27) | 0.004 | 0.17 | (0.05,0.30) | 0.007 |
| Self-esteem global score | 8 years | Child | 3796 | -0.15 | (-0.37,0.07) | 0.18 | -0.09 | (-0.34,0.17) | 0.50 |
| Self-Esteem scholastic | 8 years | Child | 3796 | -0.21 | (-0.42, -0.01) | 0.04 | -0.14 | (-0.37,0.10) | 0.26 |
| IQ measured with WISC | 8 years | Child | 3796 | -1.12 | (-1.85, -0.39) | 0.003 | -1.15 | (-1.97, -0.33) | 0.006 |
| **Outcome variable (Ordinal regression models)** | **Age** | **Report** | **Number** | **LogOdds** | **95% CIs** | **P-values** | **LogOdds** | **95% CIs** | **P-values** |
| Attention deficit hyperactivity disorder (ADHD) | 7 years | Mother and teacher | 3799 | 0.08 | (0.01,0.15) | 0.02 | 0.05 | (-0.03,0.13) | 0.23 |
| Hyperkinesis | 7 years | Mother and teacher | 3799 | 0.09 | (0.02,0.16) | 0.02 | 0.05 | (-0.03,0.13) | 0.21 |
| Conduct disorder | 7 years | Mother and teacher | 3755 | 0.05 | (-0.02,0.12) | 0.15 | 0.03 | (-0.05,0.11) | 0.46 |
| Depressive disorder | 7 years | Mother and teacher | 3751 | 0.07 | (-0.00,0.14) | 0.06 | 0.05 | (-0.03,0.13) | 0.26 |
| Generalised anxiety disorder (GAD) | 7 years | Mother and teacher | 3797 | 0.12 | (0.05,0.18) | 0.001 | 0.10 | (0.02,0.18) | 0.01 |
| Obsessive compulsive disorder (OCD) | 7 years | Mother and teacher | 3809 | 0.09 | (-0.00,0.17) | 0.06 | 0.02 | (-0.08,0.13) | 0.63 |
| Oppositional disorder | 7 years | Mother and teacher | 3789 | 0.09 | (0.02,0.15) | 0.01 | 0.07 | (-0.01,0.15) | 0.08 |
| Separation anxiety | 7 years | Mother and teacher | 3802 | 0.14 | (0.04,0.25) | 0.01 | 0.14 | (0.02,0.25) | 0.03 |
| Separation anxiety (ICD-10) | 7 years | Mother and teacher | 3802 | 0.24 | (0.10,0.38) | 0.001 | 0.21 | (0.05,0.38) | 0.01 |
| Social phobia | 7 years | Mother and teacher | 3804 | 0.11 | (0.03,0.18) | 0.01 | 0.13 | (0.04,0.21) | 0.004 |
| Specific phobias | 7 years | Mother and teacher | 3809 | 0.10 | (0.03,0.16) | 0.01 | 0.10 | (0.03,0.18) | 0.01 |
| Any disorder | 7 years | Mother and teacher | 3815 | 0.12 | (0.05,0.20) | 0.002 | 0.12 | (0.03,0.20) | 0.01 |
| Any emotional disorder | 7 years | Mother and teacher | 3815 | 0.13 | (0.06,0.20) | 0.002 | 0.13 | (0.06,0.21) | 0.001 |
| Any anxiety disorder | 7 years | Mother and teacher | 3815 | 0.12 | (0.05,0.19) | 0.001 | 0.12 | (0.04,0.20) | 0.002 |
| Any behavioural disorder | 7 years | Mother and teacher | 3789 | 0.09 | (0.02,0.16) | 0.01 | 0.08 | (0.00,0.16) | 0.05 |
| Attention deficit hyperactivity disorder (ADHD) | 10 years | Mother | 3726 | 0.08 | (0.00,0.15) | 0.04 | 0.04 | (-0.04,0.12) | 0.32 |
| Conduct disorder | 10 years | Mother | 3694 | 0.01 | (-0.06,0.08) | 0.83 | 0.01 | (-0.07,0.09) | 0.84 |
| Depressive disorder | 10 years | Mother | 3686 | 0.05 | (-0.02,0.13) | 0.13 | 0.03 | (-0.05,0.11) | 0.45 |
| Generalised anxiety disorder (GAD) | 10 years | Mother | 3723 | 0.12 | (0.05,0.19) | 0.001 | 0.11 | (0.03,0.19) | 0.01 |
| Obsessive compulsive disorder (OCD) | 10 years | Mother | 3734 | 0.14 | (0.04,0.24) | 0.005 | 0.13 | (0.03,0.24) | 0.02 |
| Oppositional disorder | 10 years | Mother | 3720 | 0.08 | (0.01,0.15) | 0.02 | 0.05 | (-0.03,0.13) | 0.21 |
| Separation anxiety | 10 years | Mother | 3597 | 0.22 | (0.12,0.32) | 1.62 x 10^-5^ | 0.22 | (0.11,0.34) | 3.27 x 10^-11^ |
| Social phobia | 10 years | Mother | 3731 | 0.11 | (0.03,0.18) | 0.006 | 0.10 | (0.01,0.18) | 0.03 |
| Specific phobias | 10 years | Mother | 3724 | 0.15 | (0.08,0.22) | 2.67 x 10^-5^ | 0.13 | (0.05,0.21) | 0.001 |
| Any disorder | 10 years | Mother | 3740 | 0.14 | (0.07,0.22) | 0.0003 | 0.11 | (0.02,0.20) | 0.02 |
| Any emotional disorder | 10 years | Mother | 3740 | 0.16 | (0.09,0.23) | 7.46 x 10^-6^ | 0.15 | (0.07,0.23) | 0.0002 |
| Any anxiety disorder | 10 years | Mother | 3740 | 0.18 | (0.11,0.25) | 4.66 x 10^-7^ | 0.17 | (0.09,0.25) | 3.11 x 10^-5^ |
| Any behavioural disorder | 10 years | Mother | 3720 | 0.08 | (0.01,0.15) | 0.02 | 0.06 | (-0.02,0.14) | 0.15 |

**Legend:** Separate linear or ordinal regressions with robust SEs adjusted for child’s sex, age, the first 5 principal components of genetic ancestry and maternal PRS for neuroticism.

### Analyses investigating whether the child PRS for neuroticism affected our outcomes of interest mainly via a genetic liability to neuroticism (and not via other pathways) by using a genome-wide significant P-value threshold

The betas are shown for a genome-wide significant (P < 5 × 10^−8^) PRS created from 113 SNPs in ALSPAC children. On the left side of the table, results from complete case analyses of the child PRS using a P < 5 × 10^−8^ threshold limited to participants with maternal genotype available but unadjusted for maternal PRS are presented. On the right side of the table, results from analyses using a child PRS at a P < 5 × 10^−8^ threshold and adjusted for maternal PRS for neuroticism are presented. Both models were adjusted for the same covariates as described in the main methods section.

### Supplementary Table 12. Associations between child PRS for neuroticism (threshold P < 5 × 10^−8^) and all psychological outcomes unadjusted and adjusted for maternal PRS for neuroticism.

|  | | | | **Complete unadjusted for maternal PRS** | | | **Complete adjusted for maternal PRS** | | |
| --- | --- | --- | --- | --- | --- | --- | --- | --- | --- |
| **Outcome variable (Linear regression models)** | **Age** | **Report** | **Number** | **Beta** | **95% CIs** | **P-values** | **Beta** | **95% CIs** | **P-values** |
| Prorated score of Carey – Activity subscale | 6 months | Mother | 4271 | 0.14 | (-0.17,0.46) | 0.369 | 0.15 | (-0.16,0.47) | 0.336 |
| Prorated score of Carey – Rhythm subscale | 6 months | Mother | 4271 | 0.24 | (-0.11,0.59) | 0.186 | 0.24 | (-0.12,0.59) | 0.187 |
| Prorated score of Carey – Approach subscale | 6 months | Mother | 4268 | 0.23 | (-0.07,0.53) | 0.132 | 0.22 | (-0.08,0.52) | 0.154 |
| Prorated score of Carey – Adaptability subscale | 6 months | Mother | 4272 | 0.22 | (-0.06,0.49) | 0.127 | 0.23 | (-0.05,0.51) | 0.102 |
| Prorated score of Carey – Intensity subscale | 6 months | Mother | 4270 | 0.08 | (-0.21,0.36) | 0.595 | 0.06 | (-0.22,0.35) | 0.657 |
| Prorated score of Carey – Mood subscale | 6 months | Mother | 4269 | 0.01 | (-0.27,0.30) | 0.924 | 0.04 | (-0.25,0.32) | 0.805 |
| Prorated score of Carey – Persistence subscale | 6 months | Mother | 4270 | -0.07 | (-0.32,0.18) | 0.588 | -0.03 | (-0.28,0.23) | 0.826 |
| Prorated score of Carey – Distract subscale | 6 months | Mother | 4271 | 0.02 | (-0.25,0.28) | 0.905 | 0.02 | (-0.25,0.29) | 0.880 |
| Prorated score of Carey – Threshold | 6 months | Mother | 4269 | -0.13 | (-0.42,0.17) | 0.411 | -0.17 | (-0.47,0.13) | 0.271 |
| Prorated score of Carey – Activity subscale | 24 months | Mother | 4241 | 0.13 | (-0.09,0.35) | 0.260 | 0.12 | (-0.11,0.34) | 0.304 |
| Prorated score of Carey – Rhythm subscale | 24 months | Mother | 4238 | 0.50 | (0.21,0.79) | 0.001 | 0.46 | (0.17,0.75) | 0.002 |
| Prorated score of Carey – Approach subscale | 24 months | Mother | 4235 | 0.003 | (-0.38,0.39) | 0.987 | -0.03 | (-0.42,0.36) | 0.871 |
| Prorated score of Carey – Adaptability subscale | 24 months | Mother | 4229 | 0.10 | (-0.11,0.31) | 0.346 | 0.08 | (-0.13,0.28) | 0.477 |
| Prorated score of Carey – Intensity subscale | 24 months | Mother | 4237 | 0.11 | (-0.11,0.34) | 0.326 | 0.11 | (-0.13,0.34) | 0.368 |
| Prorated score of Carey – Mood subscale | 24 months | Mother | 4241 | 0.08 | (-0.19,0.36) | 0.558 | 0.06 | (-0.22,0.34) | 0.673 |
| Prorated score of Carey – Persistence subscale | 24 months | Mother | 4235 | 0.13 | (-0.12,0.37) | 0.306 | 0.11 | (-0.13,0.36) | 0.354 |
| Prorated score of Carey – Distract subscale | 24 months | Mother | 4238 | -0.05 | (-0.28,0.19) | 0.696 | -0.04 | (-0.28,0.19) | 0.721 |
| Prorated score of Carey – Threshold | 24 months | Mother | 4241 | 0.16 | (-0.06,0.39) | 0.158 | 0.16 | (-0.07,0.39) | 0.166 |
| Prosocial prorated subscale of SDQ | 4 years | Mother | 3990 | -0.03 | (-0.12,0.07) | 0.613 | -0.02 | (-0.12,0.08) | 0.731 |
| Hyperactive prorated subscale of SDQ | 4 years | Mother | 3996 | 0.03 | (-0.09,0.14) | 0.664 | 0.01 | (-0.11,0.13) | 0.859 |
| Emotional prorated subscale of SDQ | 4 years | Mother | 3996 | 0.10 | (0.02,0.18) | 0.012 | 0.09 | (0.01,0.17) | 0.022 |
| Conduct prorated subscale of SDQ | 4 years | Mother | 3990 | 0.01 | (-0.06,0.08) | 0.726 | -0.002 | (-0.07,0.07) | 0.954 |
| Peer problems prorated subscale of SDQ | 4 years | Mother | 3994 | 0.03 | (-0.04,0.11) | 0.406 | 0.01 | (-0.06,0.09) | 0.732 |
| Total prorated score of SDQ | 4 years | Mother | 4061 | 0.16 | (-0.07,0.39) | 0.176 | 0.10 | (-0.13,0.34) | 0.386 |
| Prosocial prorated subscale of SDQ | 6 years | Mother | 3805 | -0.07 | (-0.16,0.02) | 0.142 | -0.07 | (-0.16,0.02) | 0.112 |
| Hyperactive prorated subscale of SDQ | 6 years | Mother | 3802 | 0.03 | (-0.10,0.15) | 0.649 | 0.02 | (-0.11,0.14) | 0.815 |
| Emotional prorated subscale of SDQ | 6 years | Mother | 3801 | 0.13 | (0.05,0.22) | 0.003 | 0.12 | (0.03,0.21) | 0.008 |
| Conduct prorated subscale of SDQ | 6 years | Mother | 3804 | 0.08 | (0.01,0.16) | 0.031 | 0.08 | (-0.00,0.15) | 0.054 |
| Peer problems prorated subscale of SDQ | 6 years | Mother | 3801 | 0.10 | (0.03,0.17) | 0.008 | 0.09 | (0.02,0.16) | 0.014 |
| Total prorated score of SDQ | 6 years | Mother | 3796 | 0.35; | (0.11,0.60) | 0.005 | 0.31 | (0.06,0.56) | 0.013 |
| Prosocial prorated subscale of SDQ | 8 years | Mother | 3668 | -0.07 | (-0.17,0.03) | 0.148 | -0.07 | (0.17,0.03) | 0.196 |
| Hyperactive prorated subscale of SDQ | 8 years | Mother | 3665 | 0.05 | (-0.08,0.18) | 0.450 | 0.03 | (-0.10,0.16) | 0.660 |
| Emotional prorated subscale of SDQ | 8 years | Mother | 3666 | 0.13; | (0.03,0.22) | 0.008 | 0.11 | (0.01,0.21) | 0.025 |
| Conduct prorated subscale of SDQ | 8 years | Mother | 3666 | 0.10 | (0.02,0.17) | 0.010 | 0.08 | (0.01,0.16) | 0.030 |
| Peer problems prorated subscale of SDQ | 8 years | Mother | 3666 | 0.06 | (-0.02,0.15) | 0.132 | 0.05 | (-0.03,0.14) | 0.221 |
| Total prorated score of SDQ | 8 years | Mother | 3663 | 0.35 | (0.08,0.62) | 0.012 | 0.28 | (0.01,0.55) | 0.045 |
| Prosocial prorated subscale of SDQ | 8 years | Teacher | 2389 | -0.05 | (-0.20,0.10) | 0.538 | -0.03 | (-0.18,0.12) | 0.700 |
| Hyperactive prorated subscale of SDQ | 8 years | Teacher | 2392 | 0.002 | (-0.16,0.16) | 0.976 | -0.02 | (-0.18,0.14) | 0.814 |
| Emotional prorated subscale of SDQ | 8 years | Teacher | 2392 | 0.14 | (0.01,0.26) | 0.031 | 0.12 | (-0.003,0.25) | 0.055 |
| Conduct prorated subscale of SDQ | 8 years | Teacher | 2391 | 0.08 | (-0.00,0.17) | 0.059 | 0.07 | (-0.01,0.16) | 0.087 |
| Peer problems prorated subscale of SDQ | 8 years | Teacher | 2392 | 0.05 | (-0.06,0.15) | 0.398 | 0.03 | (-0.07,0.14) | 0.532 |
| Total prorated score of SDQ | 8 years | Teacher | 2392 | 0.27 | (-0.06,0.61) | 0.110 | 0.22 | (-0.12,0.56) | 0.204 |
| Prosocial prorated subscale of SDQ | 9 years | Mother | 3811 | -0.04 | (-0.13,0.04) | 0.334 | -0.04 | (-0.12,0.05) | 0.383 |
| Hyperactive prorated subscale of SDQ | 9 years | Mother | 3805 | 0.02 | (-0.09,0.14) | 0.712 | 0.002 | (-0.12,0.12) | 0.976 |
| Emotional prorated subscale of SDQ | 9 years | Mother | 3802 | 0.13 | (0.04,0.21) | 0.005 | 0.11 | (0.02,0.19) | 0.018 |
| Conduct prorated subscale of SDQ | 9 years | Mother | 3806 | 0.02 | (-0.05,0.09) | 0.586 | 0.005 | (-0.07,0.08) | 0.892 |
| Peer problems prorated subscale of SDQ | 9 years | Mother | 3802 | -0.002 | (-0.08,0.08) | 0.956 | -0.02 | (-0.10,0.06) | 0.580 |
| Total prorated score of SDQ | 9 years | Mother | 3795 | 0.17 | (-0.08,0.42) | 0.188 | 0.09 | (-0.16,0.34) | 0.481 |
| Prosocial prorated subscale of SDQ | 11 years | Teacher | 2841 | -0.06 | (-0.19,0.08) | 0.408 | -0.05 | (-0.19,0.09) | 0.502 |
| Hyperactive prorated subscale of SDQ | 11 years | Teacher | 2841 | 0.09 | (-0.06,0.25) | 0.249 | 0.08 | (-0.08,0.24) | 0.307 |
| Emotional prorated subscale of SDQ | 11 years | Teacher | 2841 | 0.11 | (0.01,0.22) | 0.035 | 0.10 | (-0.00,0.21) | 0.057 |
| Conduct prorated subscale of SDQ | 11 years | Teacher | 2841 | 0.04 | (-0.05,0.13) | 0.396 | 0.04 | (-0.06,0.13) | 0.446 |
| Peer problems prorated subscale of SDQ | 11 years | Teacher | 2841 | 0.04 | (-0.07,0.15) | 0.476 | 0.03 | (-0.08,0.14) | 0.576 |
| Total prorated score of SDQ | 11 years | Teacher | 2841 | 0.29 | (-0.06,0.63) | 0.102 | 0.25 | (-0.09,0.60) | 0.150 |
| Prosocial prorated subscale of SDQ | 11 years | Mother | 3621 | -0.01 | (-0.10,0.08) | 0.795 | -0.01 | (-0.10,0.09) | 0.904 |
| Hyperactive prorated subscale of SDQ | 11 years | Mother | 3613 | 0.05 | (-0.07,0.17) | 0.421 | 0.03 | (-0.09,0.15) | 0.612 |
| Emotional prorated subscale of SDQ | 11 years | Mother | 3614 | 0.10 | (0.01,0.20) | 0.032 | 0.08 | (-0.01,0.17) | 0.095 |
| Conduct prorated subscale of SDQ | 11 years | Mother | 3622 | 0.03 | (-0.05,0.10) | 0.471 | 0.02 | (-0.06,0.09) | 0.691 |
| Peer problems prorated subscale of SDQ | 11 years | Mother | 3622 | 0.05 | (-0.04,0.13) | 0.272 | 0.03 | (-0.05,0.12) | 0.429 |
| Total prorated score of SDQ | 11 years | Mother | 3620 | 0.23 | (-0.04,0.50) | 0.088 | 0.17 | (-0.11,0.44) | 0.230 |
| Locus of control | 8 years | Child | 3796 | 0.08 | (-0.09,0.25) | 0.343 | 0.07 | (-0.10,0.25) | 0.403 |
| Self-esteem global score | 8 years | Child | 3796 | -0.14 | (-0.50,0.21) | 0.422 | -0.11 | (-0.47,0.24) | 0.526 |
| Self-Esteem scholastic | 8 years | Child | 3796 | -0.17 | (-0.49,0.16) | 0.307 | -0.13 | (-0.45,0.20) | 0.439 |
| IQ measured with WISC | 8 years | Child | 3796 | -0.58 | (-1.68,0.52) | 0.299 | -0.50 | (-1.61,0.60) | 0.373 |
| **Outcome variable (Ordinal regression models)** | **Age** | **Report** | **Number** | **LogOdds** | **Confidence Intervals** | **P-values** | **LogOdds** | **Confidence Intervals** | **P-values** |
| Attention deficit hyperactivity disorder (ADHD) | 7 years | Mother and teacher | 3799 | 0.02 | (-0.09,0.14) | 0.670 | 0.01 | (-0.11,0.12) | 0.914 |
| Hyperkinesis | 7 years | Mother and teacher | 3799 | 0.03 | (-0.08,0.14) | 0.606 | 0.01 | (-0.10,0.12) | 0.847 |
| Conduct disorder | 7 years | Mother and teacher | 3755 | 0.003 | (-0.11,0.11) | 0.955 | -0.01 | (-0.12,0.10) | 0.882 |
| Depressive disorder | 7 years | Mother and teacher | 3751 | 0.05 | (-0.05,0.16) | 0.331 | 0.04 | (-0.07,0.15) | 0.449 |
| Generalised anxiety disorder (GAD) | 7 years | Mother and teacher | 3797 | 0.02 | (-0.09,0.12) | 0.720 | 0.004 | (-0.10,0.11) | 0.936 |
| Obsessive compulsive disorder (OCD) | 7 years | Mother and teacher | 3809 | 0.10 | (-0.05,0.25) | 0.175 | 0.08 | (-0.07,0.23) | 0.310 |
| Oppositional disorder | 7 years | Mother and teacher | 3789 | 0.09 | (-0.01,0.20) | 0.087 | 0.08 | (-0.03,0.19) | 0.136 |
| Separation anxiety | 7 years | Mother and teacher | 3802 | 0.21 | (0.05,0.37) | 0.012 | 0.20 | (0.03,0.36) | 0.019 |
| Separation anxiety (ICD-10) | 7 years | Mother and teacher | 3802 | 0.39 | (0.17,0.62) | 0.001 | 0.37 | (0.14,0.59) | 0.002 |
| Social phobia | 7 years | Mother and teacher | 3804 | 0.17 | (0.05,0.29) | 0.004 | 0.17 | (0.05,0.29) | 0.005 |
| Specific phobias | 7 years | Mother and teacher | 3809 | 0.16 | (0.05,0.27) | 0.003 | 0.16 | (0.05,0.26) | 0.004 |
| Any disorder | 7 years | Mother and teacher | 3815 | 0.08 | (-0.03,0.20) | 0.150 | 0.07 | (0.04,0.19) | 0.225 |
| Any emotional disorder | 7 years | Mother and teacher | 3815 | 0.07 | (-0.03,0.17) | 0.157 | 0.06 | (-0.04,0.17) | 0.221 |
| Any anxiety disorder | 7 years | Mother and teacher | 3815 | 0.07 | (-0.03,0.18) | 0.161 | 0.06 | (-0.04,0.17) | 0.239 |
| Any behavioural disorder | 7 years | Mother and teacher | 3789 | 0.08 | (-0.04,0.19) | 0.180 | 0.07 | (-0.05,0.18) | 0.243 |
| Attention deficit hyperactivity disorder (ADHD) | 10 years | Mother | 3726 | 0.002 | (-0.11,0.12) | 0.973 | -0.02 | (-0.13,0.10) | 0.783 |
| Conduct disorder | 10 years | Mother | 3694 | 0.12 | (0.00,0.23) | 0.050 | 0.12 | (0.00,0.23) | 0.049 |
| Depressive disorder | 10 years | Mother | 3686 | 0.04 | (-0.07,0.15) | 0.507 | 0.03 | (-0.09,0.14) | 0.656 |
| Generalised anxiety disorder (GAD) | 10 years | Mother | 3723 | 0.07 | (-0.03,0.18) | 0.173 | 0.06 | (-0.05,0.17) | 0.260 |
| Obsessive compulsive disorder (OCD) | 10 years | Mother | 3734 | -0.02 | (-0.18,0.14) | 0.789 | -0.04 | (-0.19,0.12) | 0.652 |
| Oppositional disorder | 10 years | Mother | 3720 | 0.08 | (-0.03,0.18) | 0.169 | 0.06 | (-0.05,0.17) | 0.287 |
| Separation anxiety | 10 years | Mother | 3597 | 0.12 | (-0.04,0.28) | 0.134 | 0.11 | (-0.06,0.27) | 0.199 |
| Social phobia | 10 years | Mother | 3731 | 0.14 | (0.02,0.26) | 0.023 | 0.13 | (0.01,0.25) | 0.039 |
| Specific phobias | 10 years | Mother | 3724 | 0.09 | (-0.02,0.20) | 0.107 | 0.07 | (-0.04,0.18) | 0.206 |
| Any disorder | 10 years | Mother | 3740 | 0.12 | (0.001,0.23) | 0.048 | 0.10 | (-0.02,0.21) | 0.109 |
| Any emotional disorder | 10 years | Mother | 3740 | 0.09 | (-0.01,0.20) | 0.090 | 0.07 | (-0.03,0.18) | 0.173 |
| Any anxiety disorder | 10 years | Mother | 3740 | 0.18 | (0.11,0.25) | 4.66 x 10^-7^ | 0.06 | (-0.05,0.17) | 0.266 |
| Any behavioural disorder | 10 years | Mother | 3720 | 0.08 | (-0.03,0.19) | 0.150 | 0.07 | (-0.04,0.18) | 0.231 |

**Legend:** Separate linear and ordinal regression with robust SEs were employed. “Unadjusted” analyses were adjusted for child’s sex, age and the first five principal components of genetic ancestry. “Adjusted” analyses included child’s sex, age, the first five principal components of genetic ancestry and the maternal PRS for neuroticism as covariates.

### Sensitivity analyses: Addressing missing data

In this study, sample attrition on the outcome of interest was moderately high, ranging from 20% to 55%; in addition, after analysing proportions of missingness according to different socio-demographic indicators and exploring variables associated with missingness, we found evidence that the probability of having missing data depended on observed values (Missing At Random – MAR).

Therefore, we employed multiple imputation through chained equations (MICE), also known as fully conditional specification, on 100 imputed datasets with 20 cycles of regression switching for two separate cross-sectional analysis models (i.e., one model with temperament - CTSs - and one model with emotional and behavioural difficulties - SDQs - as outcomes) using the ice command in Stata 15.1 ^6–8^. Samples were restricted to those children who had genetic data and at least one outcome variable of the repeated measures available. We used the “*by*” command to perform imputations separately on boys and girls. When imputing the outcomes, we included all the variables of our models (including covariates and past and future measures of the outcome) and auxiliary variables which were associated with missingness. Imputations on the DAWBA outcome were not performed because of the low number of cases.

MICE was employed under the assumption that data were MAR. Including auxiliary variables is recommended to make the MAR assumption more plausible ^9^, however we kept the number of covariates included relatively small, selecting those with little/no missing values and that were strongly associated with levels of missingness in our models ^10^.

As we were not able to employ MI on all the analyses (e.g., DAWBA and sensitivity analysis using the maternal PRS for neuroticism) that we performed in the complete case analysis (CCA), the imputed results are presented only as a sensitivity analysis. We examined consistency across results to explore whether our complete case findings were biased because of missing data ^9^.

### Supplementary Table 13. OR for missing data in the main outcomes’ models.

|  | Mother’s educational attainment (n=12,482) | | Maternal social class (10,109) | | Maternal age at pregnancy (12,060) | | Mother’s post-partum EPDS score (12,150) | | Maternal PRS to neuroticism  (7,826) | | Child PRS to neuroticism  (7,851) | |
| --- | --- | --- | --- | --- | --- | --- | --- | --- | --- | --- | --- | --- |
| **Models** | **OR** | **95% CI** | **OR** | **95% CI** | **OR** | **95% CI** | **OR** | **95% CI** | **OR** | **95% CI** | **OR** | **95% CI** |
| Carey_6m Model^a^ | 0.65 | (0.61,0.68) | 1.52 | (1.40,1.65) | 0.93 | (0.93,0.94) | 1.05 | (1.04,1.05) | 1.07 | (1.03,1.12) | 1.10 | (1.04,1.16) |
| Carey_24m Model^a^ | 0.62 | (0.59,0.65) | 1.58 | (1.46,1.72) | 0.93 | (0.92,0.93) | 1.04 | (1.04,1.05) | 1.08 | (1.03,1.13) | 1.09 | (1.03,1.15) |
| SDQ_4y Model^b^ | 0.64 | (0.60,0.67) | 1.57 | (1.44,1.70) | 0.93 | (0.92,0.94) | 1.05 | (1.04,1.05) | 1.06 | (1.02,1.11) | 1.07 | (1.02,1.12) |
| SDQ_6y Model^b^ | 0.61 | (0.58,0.64) | 1.65 | (1.52,1.79) | 0.92 | (0.92,0.93) | 1.05 | (1.04,1.05) | 1.09 | (1.04,1.14) | 1.07 | (1.02,1.12) |
| SDQ_8y Model^b^ | 0.58 | (0.55,0.61) | 1.74 | (1.60,1.88) | 0.92 | (0.91,0.93) | 1.05 | (1.04,1.05) | 1.06 | (1.01,1.11) | 1.07 | (1.02,1.12) |
| SDQ_8y* Model^b^ | 0.79 | (0.75,0.84) | 1.22 | (1.12,1.33) | 0.96 | (0.95,0.97) | 1.03 | (1.02,1.04) | 1.01 | (0.96,1.06) | 1.03 | (0.98,1.07) |
| SDQ_9y Model^b^ | 0.59 | (0.56,0.62) | 1.78 | (1.64,1.93) | 0.92 | (0.91,0.93) | 1.05 | (1.04,1.05) | 1.05 | (1.00,1.09) | 1.07 | (1.01,1.12) |
| SDQ_11y* Model^b^ | 0.85 | (0.81,0.89) | 1.16 | (1.06,1.26) | 0.97 | (0.96,0.98) | 1.03 | (1.02,1.03) | 1.02 | (0.98,1.07) | 0.99 | (0.95,1.03) |
| SDQ_11y Model^b^ | 0.58 | (0.55,0.61) | 1.71 | (1.57,1.85) | 0.92 | (0.92,0.93) | 1.05 | (1.04,1.06) | 1.07 | (1.02,1.12) | 1.08 | (1.03,1.13) |
| DAWBA_7y Model^c^ | 0.59 | (0.57,0.63) | 1.65 | (1.52,1.78) | 0.92 | (0.92,0.93) | 1.05 | (1.04,1.05) | 1.07 | (1.02,1.11) | 1.08 | (1.03,1.13) |
| DAWBA_10y Model^c^ | 0.59 | (0.56,0.62) | 1.71 | (1.57,1.85) | 0.92 | (0.92,0.93) | 1.05 | (1.04,1.06) | 1.07 | (1.02,1.12) | 1.10 | (1.05,1.15) |

**Legend:** The models with the asterisk* represent those models where the reporter of the child emotional and behavioural problem was the teacher. Mother’s educational attainment (three levels; highest level corresponds to highest education), Maternal social class (dichotomous variable; higher levels corresponding to lower social class), Mother smoked in pregnancy (dichotomous variable (No/Yes) indicating women who smoked during the first three months of pregnancy, Mother’s post-partum depression (continuous score on EPDS for mothers of the index child). a: all Carey subscales and covariates used in the models (i.e., child’s sex, child PRS for neuroticism and the first five principal components of genetic ancestry);

b: all SDQ subscales and covariates used in the models (i.e., child’s sex, child’s age, child PRS to neuroticism and the first five principal components of genetic ancestry;

c: all DAWBA scales reported and covariates used in the models (i.e., child’s sex, child’s age, child PRS to neuroticism and the first five principal components of genetic ancestry).

### Supplementary Table 14. Associations between child PRS for neuroticism and various psychological outcomes in multiple imputed datasets.

|  |  |  | **Analyses in complete dataset** | | | | **Analysis in multiply imputed datasets** | | | |
| --- | --- | --- | --- | --- | --- | --- | --- | --- | --- | --- |
| **Outcome variable (Linear regression models)** | **Age** | **Reporter** | **N** | **Beta** | **95% Confidence Intervals** | **P-values** | **N** | **Beta** | **95% Confidence Intervals** | **P-values** |
| *Activity | 6 months | Mother | 6271 | -0.16 | (-0.33, 0.00) | 0.057 | 6871 | -0.16 | (-0.33,0.00) | 0.06 |
| *Rhythm | 6 months | Mother | 6269 | 0.08 | (-0.10, 0.26) | 0.393 | 6871 | 0.08 | (-0.10,0.27) | 0.37 |
| Approach | 6 months | Mother | 6266 | 0.18 | (0.01, 0.35) | 0.034 | 6871 | 0.18 | (0.02, 0.035) | 0.03 |
| *Adaptability subscale | 6 months | Mother | 6271 | 0.11 | (-0.04, 0.27) | 0.139 | 6871 | 0.11 | (-0.04,0.26) | 0.15 |
| *Intensity | 6 months | Mother | 6269 | 0.01 | (-0.14, 0.16) | 0.896 | 6871 | 0.01 | (-0.14,0.16) | 0.91 |
| *Mood | 6 months | Mother | 6267 | 0.03 | (-0.12, 0.19) | 0.665 | 6871 | 0.03 | (-0.12,0.18) | 0.70 |
| *Persistence subscale | 6 months | Mother | 6269 | -0.03 | (-0.16, 0.11) | 0.716 | 6871 | -0.03 | (-0.16,0.11) | 0.72 |
| *Distract | 6 months | Mother | 6271 | 0.07 | (-0.07, 0.22) | 0.323 | 6871 | 0.07 | (-0.07,0.22) | 0.32 |
| *Threshold | 6 months | Mother | 6264 | 0.21 | (0.05, 0.37) | 0.010 | 6871 | 0.21 | (0.05,0.37) | 0.01 |
| *Activity | 24 months | Mother | 6260 | 0.08 | (-0.04, 0.20) | 0.174 | 6871 | 0.09 | (-0.03, 0.21) | 0.16 |
| *Rhythm | 24 months | Mother | 6257 | 0.25 | (0.09, 0.40) | 0.002 | 6871 | 0.27 | (0.12, 0.42) | 0.001 |
| *Approach subscale | 24 months | Mother | 6253 | 0.17 | (-0.03, 0.36) | 0.092 | 6871 | 0.18 | (-0.02, 0.39) | 0.07 |
| *Adaptability subscale | 24 months | Mother | 6233 | 0.15 | (0.04, 0.27) | 0.006 | 6871 | 0.17 | (0.07, 0.28) | 0.002 |
| *Intensity | 24 months | Mother | 6255 | 0.14 | (0.02, 0.26) | 0.027 | 6871 | 0.15 | (0.03, 0.27) | 0.02 |
| *Mood | 24 months | Mother | 6259 | 0.21 | (0.06, 0.36) | 0.005 | 6871 | 0.24 | (0.09, 0.39) | 0.001 |
| *Persistence subscale | 24 months | Mother | 6252 | 0.20 | (0.07, 0.32) | 0.002 | 6871 | 0.20 | (0.08, 0.33) | 0.002 |
| *Distract | 24 months | Mother | 6255 | 0.07 | (-0.05, 0.20) | 0.260 | 6871 | 0.07 | (-0.06, 0.19) | 0.29 |
| *Threshold | 24 months | Mother | 6260 | -0.10 | (-0.21, 0.02) | 0.113 | 6871 | -0.09 | (-0.02, 0.02) | 0.11 |
| Total score of SDQ | 4 years | Mother | 5788 | 0.27 | (0.15, 0.40) | 2.30 x 10^-5^ | 7,451 | 0.30 | (0.18, 0.42) | 9.58 x 10^-7^ |
| Total score of SDQ | 6 years | Mother | 5510 | 0.37 | (0.24, 0.50) | 2.43 x 10-8 | 7,451 | 0.39 | (0.27, 0.52) | 9.64 x 10^-10^ |
| Total score of SDQ | 8 years | Mother | 5277 | 0.52 | (0.38, 0.67) | 2.08 x 10^-12^ | 7,451 | 0.53 | (0.39, 0.67) | 1.17 x 10^-13^ |
| Total score of SDQ | 8 years | Teacher | 3582 | 0.52 | (0.33, 0.71) | 8.13 x 10^-8^ | 7,451 | 0.53 | (0.35, 0.70) | 2.92 x 10^-9^ |
| Total score of SDQ | 9 years | Mother | 5516 | 0.45 | (0.31, 0.59) | 2.98 x 10^-10^ | 7,451 | 0.50 | (0.37, 0.63) | 4.76 x 10^-14^ |
| Total score of SDQ | 11 years | Mother | 5128 | 0.47 | (0.33, 0.61) | 4.70 x 10^-11^ | 7,451 | 0.48 | (0.35, 0.62) | 3.19 x 10^-12^ |
| Total score of SDQ | 11 years | Teacher | 4270 | 0.34 | (0.17, 0.51) | 8.85 x 10^-5^ | 7,451 | 0.32 | (0.15, 0.49) | 0.0002 |

**Legend**: Separate linear regressions with robust SEs adjusted for child’s sex, age and the first five principal components of genetic ancestry. *Subscales of the prorated score of the Carey Infant Temperament Scales (CTSs).

### Supplementary information on the final linear mixed effect model

**Stata code:** xtmixed logtype age_8y i.kz021_rc i.reporter i.scale_ child_3_std c.age_8y##c.chil d_3_std reporter##c.child_3_std scale_##c.child_3_std reporter##kz021_rc##c.age_8y i.scale_##i.kz021_rc##c.age_8y i.reporter##i.scale_##i.kz021_rc scale_##c.child_3_std##c.age_8y i.scale_##c.child_3_std##i.reporter c.age_8y##c.child_3_std##i.reporter mat_ed mat_sclass c994_rc pc1_child-pc5_child || ID:age_8y, cov(un) || scale:reporter, cov(un) reml

**Legend:** logtype is the natural log + 1 of the outcome (internalising and externalising scales of the SDQ); kz021 indicates the sex of the child (0=males and 1=females); child_3_std indicates the child PRS for neuroticism; age_8y indicates child’s age after having been centered at 8 years of age; reporter indicates whether the outcome was reported by the mother or by the teacher; scale indicates the subscale of the SDQ (0=Internalising and 1=Externalising); mat_ed indicates maternal education (1 indicates low education (equivalent to O level, Certificate of Secondary Education (CSE) or vocational training), 2 indicates A level, and 3 indicates some degree); Mat_sclass indicates maternal social class (0=low, 1=high); c994_rc indicates the maternal age in years during pregnancy; pc1_child-pc5_child represents the list of the first five principal components of genetic ancestry.

#### Random Effects findings

As reported in **Table 1,** at the individual child level we found strong evidence of between-child variation in both the overall levels of scores in addition to how these levels change over time. These variations were positively correlated which means that children with low scores were more stable over time while children with higher scores changed more over time. In the third level of this model, we also estimated the random variability at the scale level and its correlation with the reporter of the scale. Here, we found that the higher the difference in scores between scales (i.e., children predominantly scoring high on the externalising scale and low on the internalising scale) the lesser the impact of the reporter of the scale.

### Supplementary Table 15. Mean differences in low vs high child PRS at youngest compared to oldest child age by sex of the child and reporter.

| Mother reported | Youngest Age (4 years) | Oldest Age (11 years) | Mean difference youngest vs oldest |
| --- | --- | --- | --- |
| Females | **Mean (95% CI)** | **Mean (95% CI)** | **Mean (95% CI)** |
| Lowest PRS (SDQ Internalising) | 1.89 (1.81, 1.98) | 1.55 (1.47, 1.64) | 0.34 (0.34, 0.34) |
| Highest PRS (SDQ Internalising) | 2.15 (2.06, 2.24) | 1.94 (1.85, 2.04) | 0.21 (0.21, 0.20) |
| Mean Difference Low vs High PRS | -0.26 (-0.25, -0.26) | -0.39 (-0.38, 0.40) | 0.13 (0.13, 0.14) |
| Males | **Mean (95% CI)** | **Mean (95% CI)** | **Mean (95% CI)** |
| Lowest PRS (SDQ Internalising) | 2.04 (1.96, 2.12) | 1.47 (1.40, 1.55) | 0.57 (0.56, 0.57) |
| Highest PRS (SDQ Internalising) | 2.31 (2.22, 2.40) | 1.85 (1.76, 1.94) | 0.46 (0.46, 0.46) |
| Mean Difference Low vs High PRS | -0.27 (-0.26, -0.28) | -0.38 (-0.36, -0.39) | 0.11 (0.10, 0.11) |
| Females | **Mean (95% CI)** | **Mean (95% CI)** | **Mean (95% CI)** |
| Lowest PRS (SDQ Externalising) | 4.18 (4.04, 4.33) | 2.37 (2.27, 2.48) | 1.81 (1.77, 1.85) |
| Highest PRS (SDQ Externalising) | 4.48 (4.32, 4.64) | 2.63 (2.52, 2.75) | 1.85 (1.80, 1.89) |
| Mean Difference Low vs High PRS | -0.67 (-0.28, -0.31) | -0.26 (-0.25, -0.27) | -0.04 (-0.03, -0.04) |
| Males | **Mean (95% CI)** | **Mean (95% CI)** | **Mean (95% CI)** |
| Lowest PRS (SDQ Externalising) | 4.97 (4.80, 5.13) | 3.29 (3.16, 3.43) | 1.68 (1.64, 1.7) |
| Highest PRS (SDQ Externalising) | 5.31 (5.13, 5.49) | 3.62 (3.48, 3.77) | 1.69 (1.65, 1.72) |
| Mean Difference Low vs High PRS | -0.16 (-0.33, -0.36) | -0.33 (-0.32, -0.34) | -0.01 (-0.01, -0.02) |
| Teacher reported | **Youngest Age (8 years)** | **Oldest Age (11 years)** | **Mean difference Youngest vs Oldest** |
| Females | **Mean (95% CI)** | **Mean (95% CI)** | **Mean (95% CI)** |
| Lowest PRS (SDQ Internalising) | 1.16 (1.07, 1.25) | 1.09 (1.00, 1.18) | 0.07 (0.07, 0.07) |
| Highest PRS (SDQ Internalising) | 1.14 (1.07, 1.22) | 1.19 (1.10, 1.29) | -0.05 (-0.03, -0.07) |
| Mean Difference Low vs High PRS | 0.02 (0.0, 0.03) | -0.10 (-0.10, -0.11) | -0.08 (-0.10, 0.14) |
| Males | **Mean (95% CI)** | **Mean (95% CI)** | **Mean (95% CI)** |
| Lowest PRS (SDQ Internalising) | 1.45 (1.35, 1.55) | 1.54 (1.44, 1.65) | -0.09 (-0.09, -0.10) |
| Highest PRS (SDQ Internalising) | 1.76 (1.65, 1.87) | 1.66 (1.55, 1.78) | -0.10 (-0.10, 0.09) |
| Mean Difference Low vs High PRS | -0.31 (-0.30, -0.32) | -0.12 (-011, -0.13) | 0.19 (-0.19, -0.19) |
| Females | **Mean (95% CI)** | **Mean (95% CI)** | **Mean (95% CI)** |
| Lowest PRS (SDQ Externalising) | 1.10 (1.02, 1.19) | 0.79 (0.71, 0.87) | 0.31 (0.31, 0.32) |
| Highest PRS (SDQ Externalising) | 1.37 (1.27, 1.47) | 0.84 (0.76, 0.92) | 0.53 (0.51, 0.55) |
| Mean Difference Low vs High PRS | -0.27 (-0.25, -0.28) | -0.05 (-0.05, -0.05) | -0.22 (0.20, -0.23) |
| Males | **Mean (95% CI)** | **Mean (95% CI)** | **Mean (95% CI)** |
| Lowest PRS (SDQ Externalising) | 2.45 (2.31, 2.59) | 2.40 (2.26, 2.55) | 0.05 (0.05, 0.04) |
| Highest PRS (SDQ Externalising) | 2.89 (2.73, 3.05) | 2.50 (2.35, 2.65) | 0.39 (0.38, 0.40) |
| Mean Difference Low vs High PRS | -0.44 (-0.42, -0.46) | -0.10 (-0.09, -0.10) | -0.34 (-0.33, -0.36) |

### Supplementary Figure 1. Graphical representation of potential dependent differential misclassification.

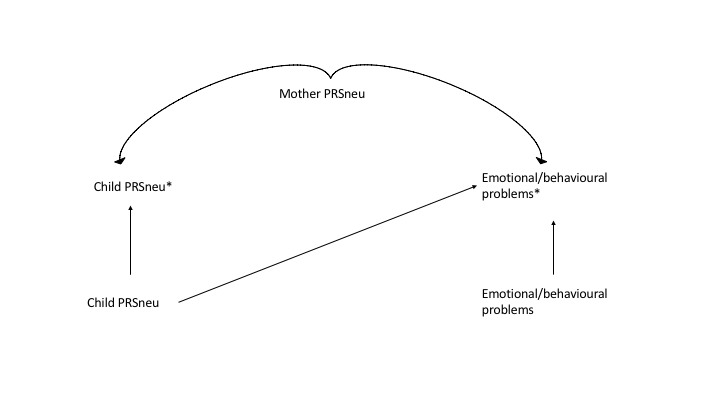

**Legend:** The variables with the asterisk* represent the measured exposure and the measured outcome. Maternal PRS for neuroticism is a variable leading to correlated/dependent measurement errors: lower versus higher levels of maternal PRS for neuroticism could differentially impact measurement of the child’s emotional and behavioural problems.

### Supplementary Figure 2. Flow diagram of ALSPAC participants with genotype information and main outcomes.

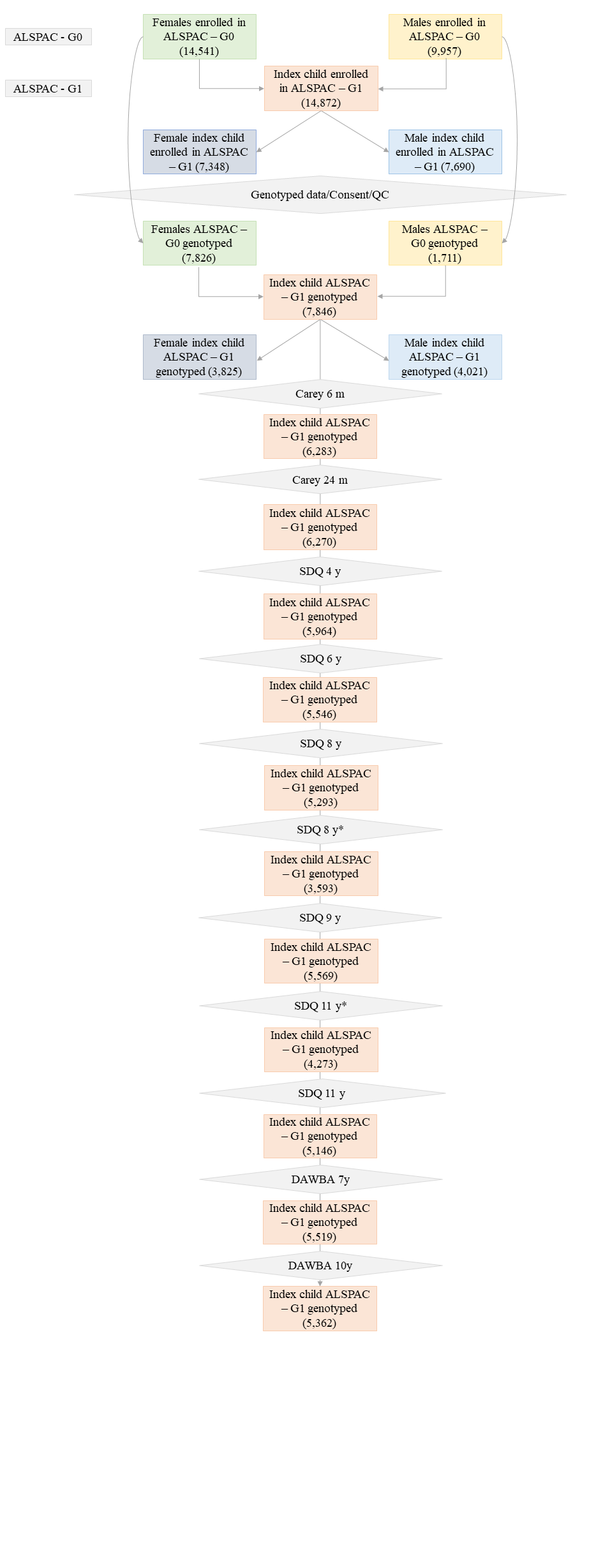

### Supplementary Figure 3. Directed Acyclic Graph (DAG) representing potential pathways through which children behave/experience emotional symptoms differently in presence of a parent.

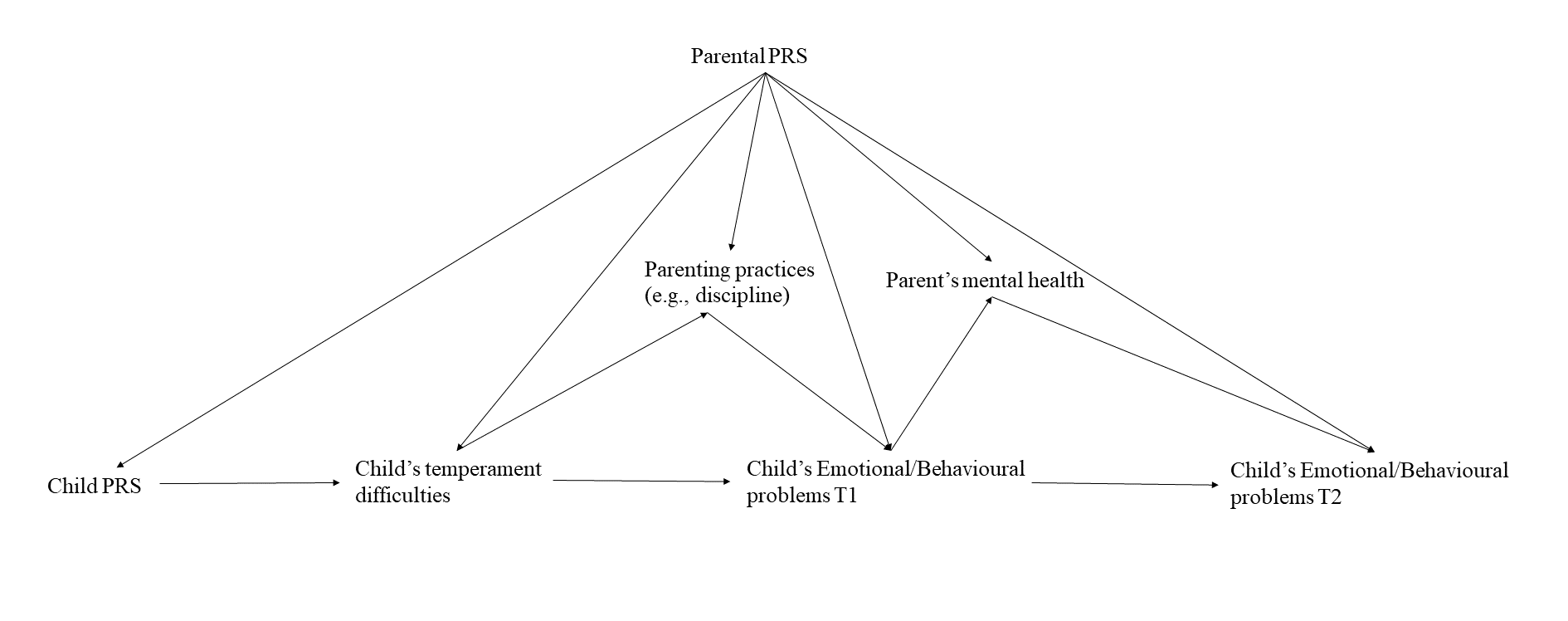

**Legend:** This DAG illustrates a hypothesised causal pathway from a child PRS for neuroticism to temperamental, emotional and behavioural problems and its relationship with parental genotype, parental parenting practices and parental mental health.
